# Enhancing, controlling, and sterilizing dengue immunity and the development of broadly protective responses

**DOI:** 10.64898/2026.03.26.26349424

**Authors:** Camila D. Odio, Rosemary A. Aogo, Saba Firdous, Charlie Voirin, Silvia Blanco-Rivera, Kelsey E. Lowman, Nhyira Asante, Ysabelle Broderson, Keerthi Konda, Chloe M. Hasund, Amparo Martinez-Perez, Patrick I. Mpingabo, Melissa Law, Caitlin Jarvis, Viviane Callier, Sally Hunsberger, Maria Abad Fernandez, Aravinda de Silva, Robbie Kattappuram, Gitanjali Bhushan, Christina Yek, Jessica Manning, Anna P. Durbin, Jeffrey I. Cohen, Daniela Weiskopf, Stephen S. Whitehead, Leah C. Katzelnick

## Abstract

Dengue is the canonical viral disease for which immune history predicts protective versus pathogenic responses and immunogenicity. Yet, due to limitations in animal models and clinical presentation after peak viremia, how pre-infection and early immune responses affect dengue outcomes is not confirmed. We conducted a phase 1 clinical trial with 45 healthy adults to test if secondary infection challenge with a heterotypic, full-length, attenuated virus increases viremia and immunogenicity compared to primary and tertiary infection. Viremia was associated with more, but still mild, clinical signs and symptoms, and secondary infection predicted greater viremia and neutralizing antibodies. However, those with the highest baseline enhancing antibodies experienced delayed inflammatory and adaptive activation, the highest viremia, strong acute immune responses, but waning of potent CD8^+^ T cells and antibodies. Baseline antibodies to non-structural protein 1 of multiple serotypes predicted early interferon and balanced immune activation, viremia control, and development of enduring, potent B and T cells, revealing how vaccines can induce broad long-lasting protection. Finally, these antibody and T cell profiles at baseline predicted sterilization of infection. We demonstrate that controlled human challenge can delineate coordinated versus dysregulated acute responses and effects on immunogenicity, informing therapeutic and vaccine strategies for dengue and other viral diseases.

## Main

Immune memory enables the body to respond to related pathogens, requiring a carefully coordinated response to control infection without excess inflammation, which can itself cause disease. Because most human pathogens with antigenically related subtypes are either emerging and locally restricted or endemic globally, direct comparisons of those with no, one, or multiple prior exposures are difficult. The four dengue virus serotypes (DENV1-4) are mosquito-borne members of the *orthoflavivirus* genus with increasing burdens but still limited ranges, causing an estimated 50 million dengue cases annually^1^. Unlike for most pathogens, viremia and disease severity are greatest during a second infection with a different DENV serotype but drop for subsequent heterotypic infections^2, 3^. Pre-existing cross-reactive antibodies to DENV can increase infection of a new serotype via Fcγ receptor-mediated phagocytosis, and such antibodies are associated with higher viremia and disease risk, including fluid leak, shock, hemorrhage, organ failure, and death^4, 5, 6, 7^. However, a hallmark of the post-secondary response is the development of broadly protective anti-DENV antibodies and T cells^8, 9, 10, 11^. These unique features make DENV useful for characterizing how immune imprinting affects responses to each sequential exposure^12, 13^ and potentially also for understanding immune dysregulation, which for other diseases may be due to intrinsic host factors that are difficult to predict and control^14^.

A major challenge to preventing and treating dengue is inducing immunity sufficient to protect individuals against each of the four serotypes, bypassing induction of an immune response that increases disease. While dengue vaccine candidates aim for tetravalent protection, the first licensed dengue vaccine increased risk of hospitalization in those who had never been infected with DENV^15^. Other dengue vaccine candidates have different designs and better efficacy and safety profiles^16, 17^, and some induce antibodies and CD8^+^ T cells associated with post-secondary protection^18, 19^. Natural infection, animal, and vaccine studies have identified correlates of protection against dengue^20, 21, 22, 23^, but no study has been designed to identify the features of sterilizing and controlling versus enhancing immunity. Further, given limited controlled human studies in DENV immune participants at risk of enhancement^24^, it is also not known how viremia and the early acute response affect later inflammatory and adaptive immunity.

## Results

Here, we conducted a controlled challenge with a live-attenuated monovalent DENV3 vaccine candidate in 45 healthy adults with no, one heterotypic, or multiple prior DENV infections (NCT05691530^25^). Strain rDEN3Δ30/31-7164 contains all viral proteins of wild-type DENV3, with attenuating mutations in non-coding regions. Participant groups were defined by antibodies measured at day 0 and medical history, including those: without evidence of flavivirus infection (‘naïve’, n=14), with neutralizing antibodies consistent with primary infection with DENV1, DENV2, or DENV4 (‘heterotypic’, n=18), and with evidence of multiple prior DENV infections, defined as balanced neutralizing antibodies to multiple serotypes (<4-fold difference in titers) and the highest titer to a serotype other than DENV3 (‘polytypic’, n=13) (**Extended Data Figs. 1a,b, Supplementary Table 1**). There were no significant differences in sex, age, race, or ethnicity between groups (**Extended Data Table 1a**). Among those with DENV immunity, 22 reported a prior dengue case, which occurred a median of 13.5 years before vaccination (**Supplementary Table 2**). All participants were subcutaneously inoculated with rDEN3Δ30/31-7164 and followed closely after vaccination. Samples were collected at days 0, 1, 3, 6, 9, 12, 15, 28, 57, 90, 180, and 365 (**Extended Data Fig. 1c**). Participants, clinical staff, and laboratory personnel were blinded to DENV histories until all participants completed through day 57.

Our trial met our primary endpoints of safety, vaccine viremia, and immunogenicity. The virus was safe and well-tolerated among all recipients (**Supplementary Table 3 and 4, Supplementary Data File 1**). As hypothesized, mean peak viral RNAemia (hereafter, viremia) of the vaccine strain was significantly higher in heterotypic than the naïve individuals (**Fig. 1a, Extended Data Fig. 2a**), consistent with enhancement of infection. All immune groups had significant increases in the geometric mean of neutralizing antibody titers to DENV1-4 at days 28 and 57 after challenge, but the largest increases occurred in the heterotypic group (**Fig. 1b, Extended Data Fig. 2b,c**). Together, these findings suggest heterotypic immunity was associated with both increased viremia and immunogenicity.

**Fig. 1.**
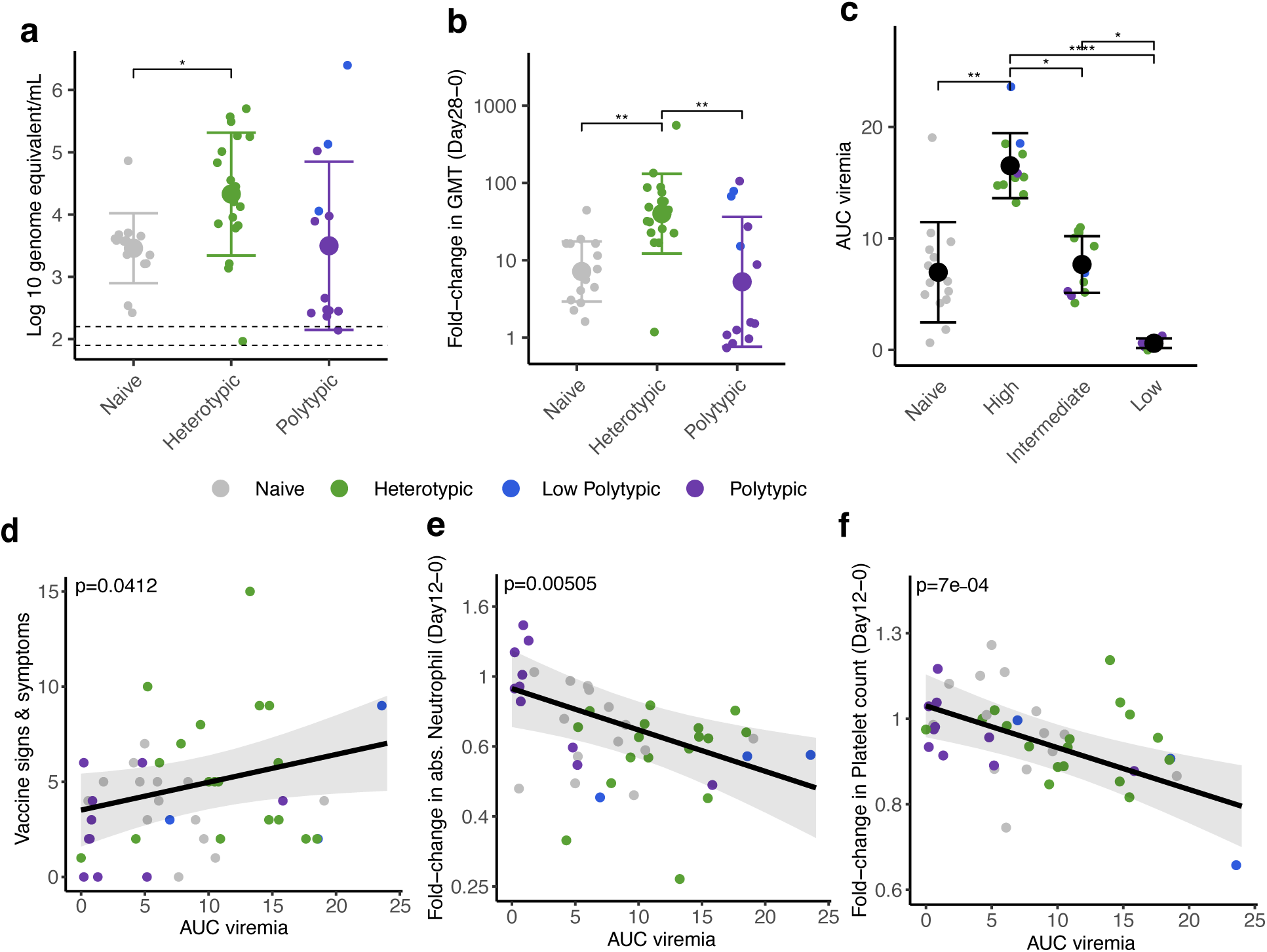
Heterotypic immunity is associated with higher viremia and greater immunogenicity, and viremia is associated with both symptom burden and laboratory changes. (a) Peak viremia, measured as RNA genome equivalent per mL on days 3, 6, 9, 12, and 15 post vaccination. As pre-specified, immune groups were compared using t-tests with Bonferroni correction. Dotted lines represent lower limit of detection (LLOD) and ½ LLOD respectively. (b) Fold-change in the geometric mean of neutralizing antibodies to standard (partially immature) DENV1-4 from day 0 to 28 by immune group. (c) The area under the curve (AUC) of viremia for immune participants was clustered using partitioning around medoids into three groups. Naïve individuals are grouped separately. For b and c, group comparisons were evaluated by Kruskal-Wallis test followed by Dunn’s test with a Bonferroni correction for pairwise comparisons. All individual data points are plotted, and group mean and standard deviations shown. * p<0.05, ** p<0.01, *** p<0.001, **** p<0.0001. Linear regression models adjusted for demographic covariates were used to evaluate associations between AUC viremia and (d) total number of vaccine-related adverse events or fold-change between day 0 and 12 in (e) absolute neutrophil or (f) platelet count. Points are colored by immune groups, with the polytypic group with low but balanced titers (low polytypic) colored separately.

To evaluate if higher viremia was associated with better post-vaccination immune responses, we stratified the immune participants based on the area under the curve (AUC) of their viremia (**Fig. 1c, Extended Data Fig. 2d, Extended Data Table 1b)**. AUC viremia was significantly associated with established markers of DENV infection, including symptom burden and reductions in absolute neutrophil and platelet counts, supporting its clinical relevance (**Fig. 1d-f**). Using unbiased clustering to analyze AUC viremia, we identified groups with almost no viremia (low, n=8), with viremia of similar magnitude to the naïve group (intermediate, n=12), and with significantly greater viremia than other groups (high, n=11). Most participants with high and intermediate viremia had heterotypic immunity (73% and 75%), while most with the low viremia had polytypic immunity (88%). We then evaluated the expansion of potent antibodies and T cells following vaccination using an immunofocus reduction neutralization assay with mature DENV1-4 virions and an activation-induced marker (AIM) T cell assay (**Supplementary Fig. 1**). Those with low viremia had minimal changes in antibodies and T cell populations post-vaccination, consistent with sterilizing immunity (**Fig. 2a-c, Extended Data Figs. 2e,f, 3 and 4**). Across timepoints, activated CD4^+^ T cells were similarly expanded in the naïve, intermediate and high viremia groups (**Fig. 2a, Extended Data Fig. 3).** At day 15, activated CD8^+^ T cells were similarly elevated in participants with high and intermediate viremia (**Fig. 2b**). However, by day 28 and 90, those with intermediate viremia had the greatest expansion of DENV-specific CD8^+^ cells, especially T effector memory re-expressing CD45RA cells (Temra), which have previously been associated with protective post-secondary immunity^10^ (**Fig. 2b,c, Extended Data Fig. 4**). In contrast, those with high viremia had low magnitude CD8^+^ Temra and other memory subsets, similar to naïve individuals. Further, those with high viremia experienced rapid decay of neutralizing antibodies to all four mature DENV serotypes than those with intermediate viremia, even out to 365 days post-vaccination (half-life of 1.2 vs. 2.9 years for intermediate, p<0.05) (**Fig. 2d-f, Extended Data Fig. 5**). Neutralization of mature DENV virions has previously been identified as a more sensitive measure of virus-specific and broad protective antibodies^11, 26, 27, 28^_._

**Fig. 2.**
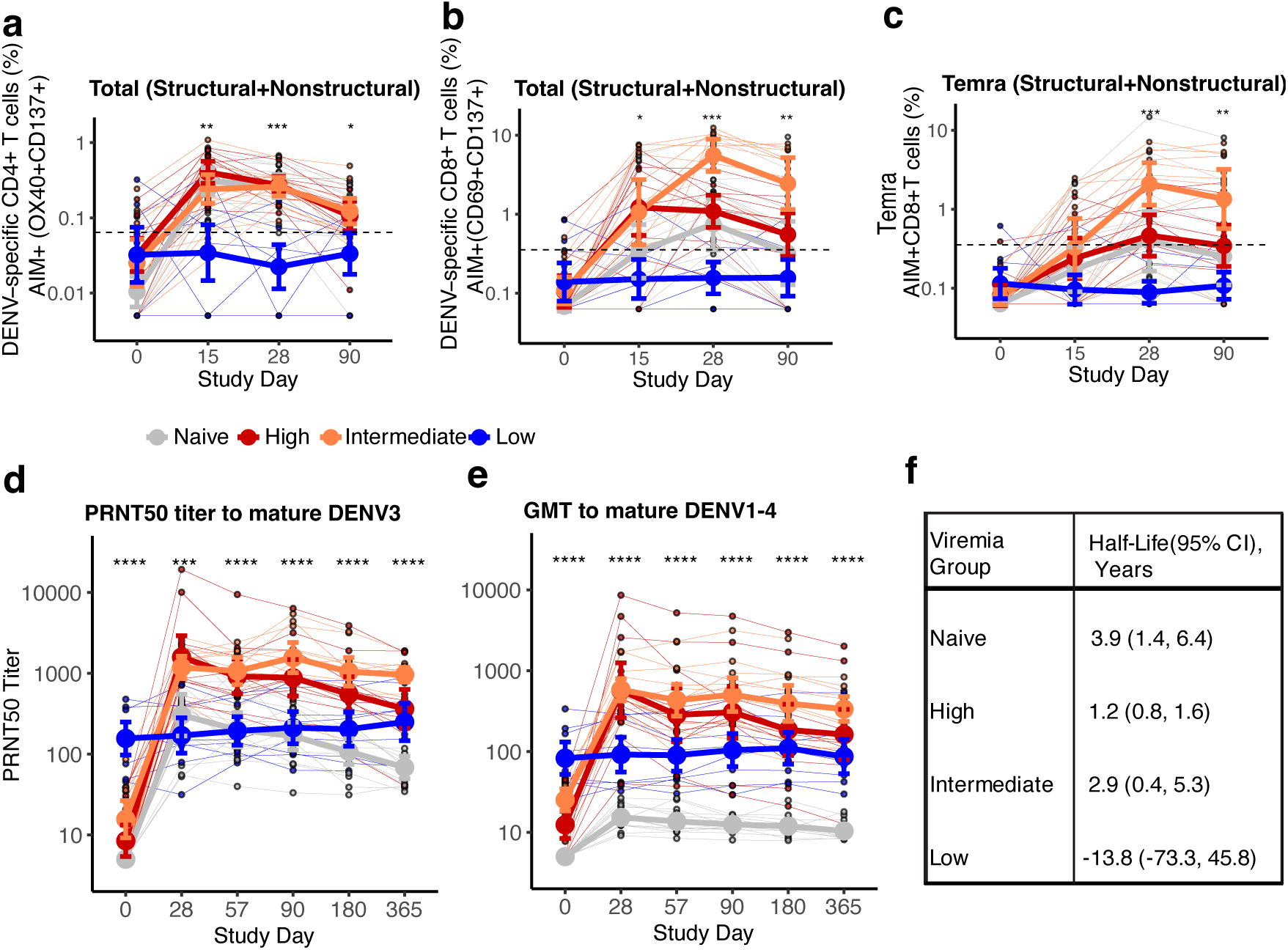
**Those with high viremia have less potent and enduring adaptive responses than those with intermediate viremia**. Total magnitude of DENV-reactive CD4^+^ (**a**), CD8^+^ (**b**) and CD8^+^ T effector memory re-expressing CD45RA, or Temra (**c**) measured at day 0, 15, 28, and 90 by activation induced marker (AIM) assay, colored by viremia group. AIM^+^ CD4^+^ T cells were defined as OX40^+^CD137^+^ and AIM^+^ CD8^+^ T cells by CD69^+^CD137^+^. The dotted line shows the limit of sensitivity. Neutralizing antibody titers to mature DENV3 (**d**) or the geometric mean of neutralizing antibody titers (GMT) to mature DENV1-4 (**e**) measured at each timepoint by immunofocus reduction neutralization test, colored by viremia group. (**f**) Half-life and 95% confidence interval (CI) of the neutralizing antibody GMT to mature DENV1-4 for each viremia group, estimated using a linear mixed-effects model with time-by-group interaction and a subject-specific random intercept to account for repeated measures within individuals. All group comparisons were evaluated by Kruskal-Wallis test. All individual data points are plotted, and group mean and standard deviations shown. * p<0.05, ** p<0.01, *** p<0.001, **** p<0.0001.

Those with intermediate viremia may have had better existing humoral and cellular immunity than those with high viremia and were able to refine this memory during the acute secondary response. We measured pre-challenge antibodies with binding and function assays, including those that have shown promise as correlates of protection, as well as more commonly used assays (**Extended Data Figs. S6-7, Supplementary Fig. 2**). T cells were measured by AIM assay with intracellular cytokine staining, although few had detectable baseline responses. We used multinomial elastic net regression to identify which of the 80 distinct immune measures best discriminated between viremia responses; similar results were observed for linear models of AUC viremia and with random forest models (**Supplementary Fig. 3**). We found that low viremia was predicted by measures of potent neutralizing antibodies and targeting conserved quaternary structure epitopes, including neutralization of mature DENV3 virions, competition of serum with a broadly neutralizing antibody for binding to the DENV3 envelope dimer epitope, and neutralization of multiple DENV serotypes (**Fig. 3a**). Both those with intermediate and high viremia had some enhancing antibodies and lower mature neutralizing antibodies (**Fig. 3b**). However, intermediate viremia was associated with binding antibodies to non-structural protein 1 (NS1) of multiple DENV serotypes and living in dengue-endemic areas more recently (**Fig. 3a** and **c**). High viremia was best predicted by enhancing antibodies measured in minimally diluted serum on a monocytic cell line (U937) engineered to express all human Fcγ receptors (CD32, CD64, CD16).

**Fig. 3.**
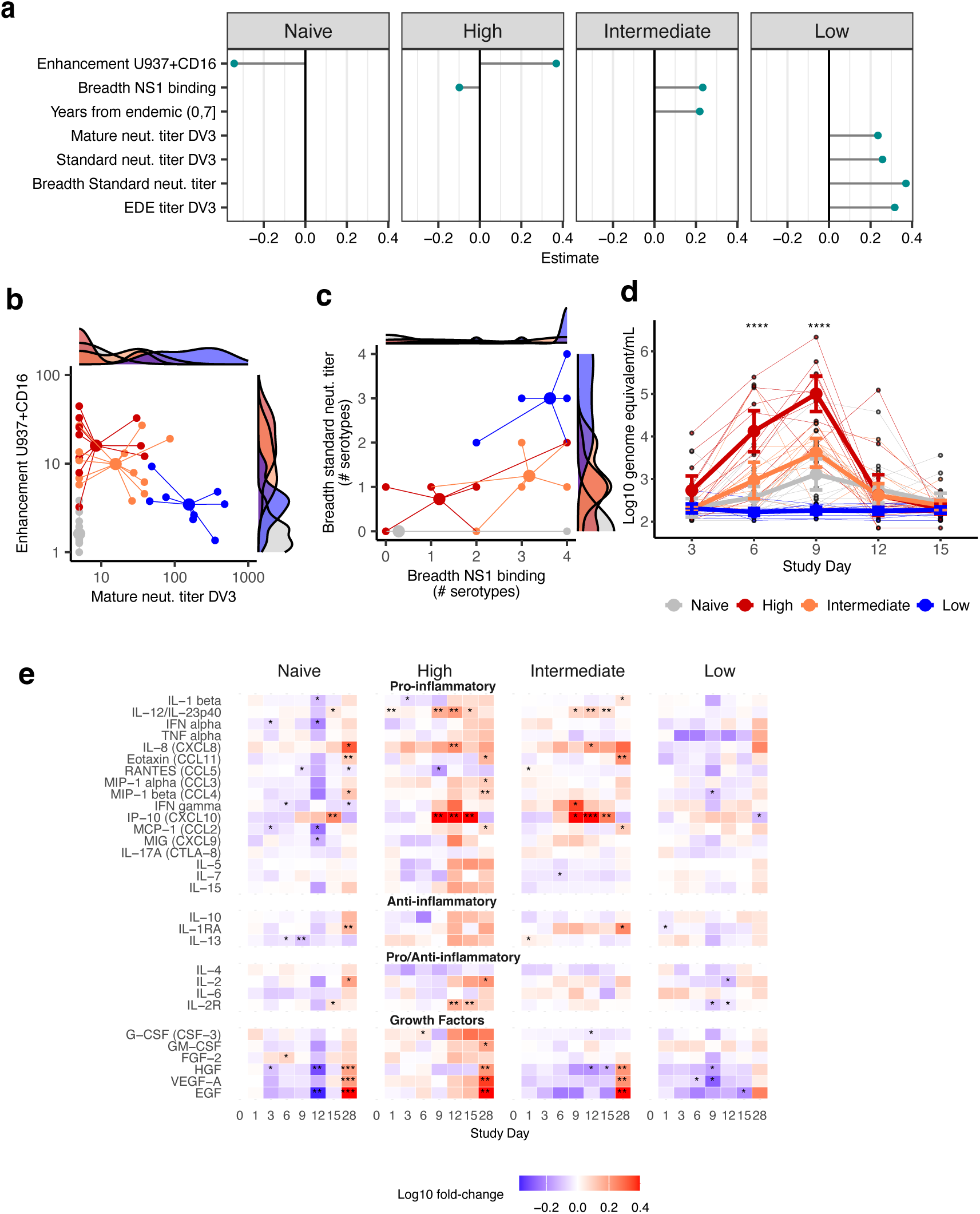
Baseline antibody-dependent enhancement is associated with delayed, broader inflammatory responses. (**a**) Results of multinomial elastic net regression showing the best predictors of viremia groups. Coefficients with effects >|0.2| are shown. Positive coefficients indicate that a higher magnitude response predicted being in that group. Star plots of (**b**) neutralizing antibody titers to mature DENV3 versus fold-enhancement of DENV3 infection by immune sera at a 1:10 dilution on U937+CD16 cells and (**c**) the number of DENV serotypes to which each individual had NS1 positivity versus the number of standard DENV serotypes neutralized at a 1:60 dilution. Lines connect datapoints for each viremia group to the estimated group mean, and marginal distributions are shown. (**d**) Kinetics of viremia, measured as RNA genome equivalent per mL, at each timepoint by viremia group. Group comparisons were evaluated by Kruskal-Wallis test. Individual data points are plotted, and group mean and standard deviations shown. (**e**) Heatmap of the average log-fold change from day 0 at each timepoint for cytokines, chemokines, and growth factors, separated by viremia group. Fold-change from day 0 within each group for each cytokine was evaluated by Wilcoxon signed-rank test. * p<0.05; ** p<0.01; *** p<0.001; **** p<0.0001.

*In vitro* and animal studies show entry into immune cells via antibody-dependent enhancement (ADE) inhibits intracellular antiviral responses and produces aberrant immune cell signaling, enabling the virus to replicate more without triggering a normal innate and adaptive response^29, 30, 31^. Consistent with ADE, the high viremia group had elevated viremia at day 3 and 6 and higher peak viremia at day 9 (**Fig. 3d**). To evaluate the systemic response, we measured clinical outcomes, biomarkers, serum cytokines, chemokines, and growth factors from days 0 through 28. There were modest but not significant differences in total vaccine-associated signs and symptoms across groups, and naïve, intermediate, and high viremia groups experienced a drop in white blood cell counts during and after peak viremia (**Extended Data Fig. 8**). Those with low viremia had minimal changes in cell counts or cytokines, consistent with a sterilizing response (**Fig. 3e**). Both those with intermediate and high viremia had similar, very mild decreases in platelets and elevations of C reactive protein and liver enzymes, suggesting their clinical presentations were not significantly different. However, inflammatory responses were remarkably different. High viremia was associated with early increases in tissue and endothelium activation (G-CSF, VEG-F), but without an interferon response (IFN-α or IFN-γ), suggestive of early viral dissemination and attenuated activation of innate immune pathways. Yet, after peak viremia, and through day 28, those with high viremia had broad elevation of 25/30 cytokines tested, including those observed in severe secondary dengue and associated with leak (IL-10, TNF-alpha, IL1-β). In contrast, the intermediate viremia group had an early increase in antiviral-associated cytokines (IFN-α, IFN-γ, IP-10) with a moderating anti-inflammatory response (IL-1RA), followed by a limited inflammatory response associated with protective immunity (IL-12, IL-8, IP-10), including markers also seen in naïve individuals.

To evaluate if the high viremia group also experienced a delayed, then rapid adaptive response, we looked at changes in DENV-reactive IgG antibodies by ELISA, IgG^+^, IgA^+^, and IgM^+^ plasmablasts by ELISpot, and T cells by AIM assay during the acute phase. At day 9, only the intermediate group had detectable DENV3-reactive IgG^+^ plasmablasts, suggesting an earlier response (**Fig. 4a**). DENV3-specific antibodies and IgG^+^ plasmablasts reactive to DENV3 and other serotypes increased between days 9, 12, and 15 in those with intermediate and high viremia, but the expansion was faster and the final magnitude greater in those with high viremia (**Fig. 4a-d**). A similar pattern was observed for IgA^+^ plasmablasts, and none of the groups had significant IgM^+^ plasmablasts (**Extended Data Fig. 9a-c**). By day 28 and 57, there were no differences in binding antibodies between intermediate and high viremia groups, and both developed potent cross-reactive neutralizing antibodies to mature DENV and targeting the envelope dimer epitope of multiple serotypes, populations not observed in naïve individuals (**Fig. 2d-f** and **4d-f**, **Extended Data Fig. 5** and **9d-g**). The finding of more gradual B cell expansion in the intermediate group mirrored the earlier increase in serum IFN-γ in the intermediate (day 6) than high viremia group (day 9) (**Fig. 3e**), which during secondary infection is mostly produced by T cells. We thus tested if the kinetics of IFN-γ^+^ T cells also differed. IFN-γ^+^ CD4^+^ and CD8^+^ T cells were similar across groups at day 15, if anything elevated in the high viremia group (**Fig. 4g-i)**. However, only the high viremia group had a rapid decline in IFN-γ^+^ activated T cells by days 28 and 90. A delayed type I IFN response in this group may have impaired survival of activated CD8^+^ T cells and subsequent expansion of IFN-γ^+^ and Temra cells, as observed during acute severe dengue^32^. In contrast, the intermediate group had their peak in IFN-γ^+^ cells at day 28, especially to non-structural protein epitopes, suggesting a more gradual but enduring response across the polyprotein, which may have contributed to their development of robust long-lived immunity.

**Fig. 4.**
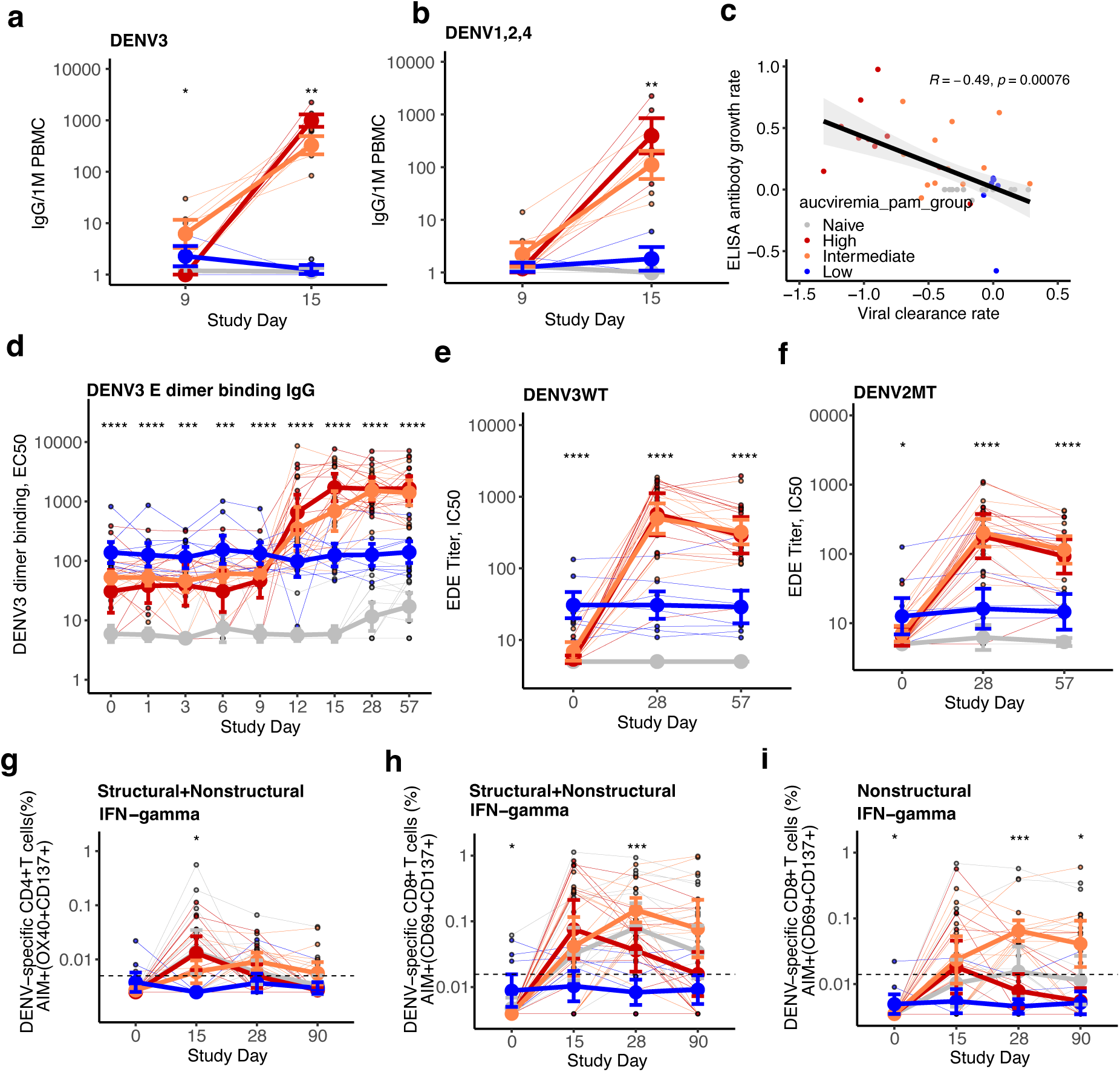
High viremia was associated with delayed, then rapid adaptive activation, whereas those with intermediate viremia had earlier, more gradual adaptive responses. IgG^+^ plasmablasts per one million (1M) peripheral blood mononuclear cells (PBMCs) that were reactive to (**a**) DENV3 and (**b**) DENV1, DENV2, and DENV4, evaluated in freshly collected samples by ELISpot. A random subset of each viremia group was tested: n=5 naïve, n=5 high, n=6 intermediate, and n=3 low. (**c**) Spearman’s correlation and p-value of the rates of change from day 9 to 12 in viremia versus binding antibodies to DENV3 E dimer measured by IgG ELISA for all participants. The rate of viral clearance and rate of antibody growth were quantified by the slope of the log-transformed change between the two timepoints. (**d**) Kinetics of DENV3 E dimer binding IgG antibodies measured by ELISA for all participants from days 0 to 57. Serum competition with envelope dimer-epitope antibody C8 for binding to the (**e**) DENV3 dimer (DENV3WT) and (**f**) DENV2 dimer with a mutation to reduce binding to the fusion loop (DENV2MT) at days 0, 28, and 57. Magnitude of IFN-γ^+^ AIM^+^ (**g**) CD4^+^ cells and (**h**) CD8^+^ T cells, and (**i**) IFN-γ^+^ AIM^+^ CD8^+^ T cells targeting only non-structural protein epitopes. For all plots, differences across groups were evaluated by Kruskal-Wallis test. Individual data points are plotted, and group mean and standard deviations shown. * p<0.05; ** p<0.01; *** p<0.001; ****p<0.0001.

## Discussion

Collectively, we show rapid activation of adaptive effector functions required to control an unchecked viral infection due to ADE came at the expense of developing the highest quality convalescent adaptive response. Those who better controlled viremia had more coordinated early innate and adaptive activation that enabled them to moderate viral replication while simultaneously developing high-quality, enduring adaptive immunity. Potent pre-existing neutralizing antibodies mediated sterilizing protection, which limited infection and resulted in minimal evolution of the adaptive response.

We show ADE on a cell line including CD16, the Fcγ receptor sensitive to afusoylated antibodies, was significantly predictive of viremia, integrating previous findings that greater afucosylated antibodies are observed during acute severe dengue and that ADE in minimally diluted serum on fresh human monocytes is associated with disease^33, 34, 35^. Further, we showed ADE, even during mild infection, hampers early anti-viral responses and was associated with delayed but broad and sustained inflammation. During natural infection, ADE likely directly and indirectly increases early endothelial barrier disruption that can become catastrophic following aberrant adaptive activation^7^, characterized by dysregulated cytokine signaling, reduced antigen presentation, pro-inflammatory NK cells, cross-reactive CD8^+^ T cells with weak killing activity, and rapid expansion of DENV-specific plasmablasts^36, 37, 38, 39^. This threshold-driven escalation of disease mirrors severe COVID-19 and influenza; once early damage surpasses a critical tipping point, rising viral burden and T cell-mediated cytotoxicity exacerbate tissue and vascular injury to the point where patients cannot recover lung function^40, 41^. In contrast, those with intermediate viremia had signatures of early immune regulation, supporting the use of immunomodulators, potentially paired with antivirals, soon after presentation. Such drugs will soon be evaluated for preventing progression to severe dengue in phase 3 trials and have been used to treat COVID-19^42, 43^.

We observed sterilizing immunity in those with potent, quaternary antibodies, highlighting the importance of vaccines inducing such antibodies to prevent viral infection and transmission^18, 44^.

In those with enhancing antibodies and lacking neutralizing antibodies, NS1 antibodies and living and in dengue-endemic areas more recently were associated with partial viral control. These measures may mark recent or multiple DENV exposures, better cellular memory across the polyprotein, or prevention of NS1-mediated vascular permeability and viral dissemination^23, 45^. This finding suggests dengue and other vaccines that induce humoral and possibly cellular immunity to non-structural proteins may not reduce viremia but may prevent severe disease^46^. Further, gradual development of adaptive immunity may result in optimal memory expansion following subsequent natural infection, similar to how slow delivery immunization induces better neutralizing but not more overall antibodies than bolus immunization^47^. Finally, the presence of enhancing antibodies measured with relevant cell lines but without moderating protective humoral or cellular immunity may be an indicator of vaccine safety risk^15^, and may imprint individuals to develop less protective immunity after subsequent infection. Identification of the features of immune memory associated with acute inflammatory outcomes as well as enduring protective immunity in a controlled human study informs therapeutic treatment and development of more broadly immunogenic vaccines.

## Data Availability

Deidentified immunological and clinical data will be deposited at ImmPort. Additionally, all data required for reproducing the figures and associated code were managed in GitHub and a copy of the code is available through Zenodo: 10.5281/zenodo.19238788.

https://zenodo.org/records/19238788

## Acknowledgments

We sincerely thank the study participants. We also thank the NIH Clinical Center staff in pharmacy, OP8, and the SCSU, Douglas Fritz and Emerito Amaro-Carambot for support with qRT-PCR assay optimization, Luz Fuentes and Douglas Kuhns for support with sample processing, Admassu Dareskedar and Melanie Cohen for support with cytokine data acquisition, and Heidi Hall for support designing the REDCAP exposure history surveys.

## Funding

Intramural Research Program of the National Institutes of Health (LCK, SH, JC, SSW, CDO) National Institutes of Health Bench-to-Bedside Program Funds Award #994875 (CDO, LCK) National Institutes of Health Director’s Challenge Innovation Award (RAA, LCK)

National Institutes of Health ReVAMPP grant U19AI181960-01 (AdS) National Institutes of Health P01AI106695 (AdS, DW) National Cancer Institute, NIIH Contract No. 75N91019D00024 (ML, CJ, VC) The contributions of the NIH authors were made as part of their official duties as NIH federal employees, are in compliance with agency policy requirements, and are considered Works of the United States Government. The findings and conclusions described here are those of the authors and do not necessarily represent the views of the NIH or the U.S. Department of Health and Human Services. In relation to Contract No. 75N91019D00024, the content of this publication does not necessarily reflect the views or policies of the Department of Health and Human Services, nor does mention of trade names, commercial products, or organizations imply endorsement by the U.S. Government.

## Author contributions

Conceptualization: CDO, RAA, KEL, APD, SH, SSW, LCK. Methodology: CDO, RAA, SF, CV, SBR, KEL, NA, YB, KK, CMH, AMP, PIM, ML, RK, MAF, AdS, JIC, DW, APD, SSW, LCK. Investigation: CDO, RAA, SF, CV, SBR, KEL, NA, YB, KK, CMH, AMP, PIM, ML, CJ, VC, MAF, AdS, GB, JIC, DW, LCK. Visualization: CDO, RAA, NA, KK, YB, LCK. Funding acquisition: SH, JC, SSW, CDO, RAA, LCK. Project administration: CDO, RAA, SBR, CJ, ML, VC, CY, JM, JIC, SSW, LCK. Supervision: LCK, RK, CDO, RAA, SH, CY, JM, JIC. Writing –original draft: LCK, RAA, CDO. Writing – review & editing: all.

## Competing interests

JM is currently an employee of Sanofi Pasteur. DW has been a consultant for Moderna and filed for patent protection for various aspects of T cell epitope and vaccine design work. AdS is listed as an inventor on pending patent applications filed by the University of North Carolina related to flavivirus vaccines and diagnostics.

## Supplementary Information is available for this paper

**Data and materials availability:** Deidentified immunological and clinical data will be deposited at ImmPort. Additionally, all data required for reproducing the figures and associated code were managed in GitHub and a copy of the code is available through Zenodo: 10.5281/zenodo.19238788. This study did not generate new unique reagents. Correspondence and requests for materials should be directed to LCK and CDO.

## Materials and Methods

### Ethical statement

This study was approved by the National Institutes of Health Institutional Review Board (NCT05691530)^25^, and all participants signed written informed consents.

### Study aims

The aims of our study were to: 1) evaluate how infection history influences vaccine safety and immunogenicity, 2) test whether infection history and immune characteristics influence protection against viremia and symptoms, 3) evaluate whether the vaccine activates distinct T cells, memory B cells, and innate cells by infection history, 4) evaluate the magnitude, durability, and breadth of protective immunity.

### Primary and secondary objectives and endpoints

Our primary objective was to evaluate the safety of monovalent DENV3 vaccination in those with distinct natural DENV infection histories living in non-endemic areas, and how prior DENV immunity influences protection against vaccine strain infection evaluated by the change in geometric mean neutralizing antibody titers and mean peak viremia. Our endpoints were: 1) The frequency and severity of local and systemic reactogenicity signs and symptoms during the 28-day period after each vaccination, unexpected adverse events (AEs) up to 28 days after each vaccination, and serious adverse events (SAEs) through day 180; 2) Change in DENV neutralizing antibody geometric mean titer (GMT) between days 0 and 28; and 3) Mean peak viremia among groups as measured by viral quantitative reverse transcription polymerase chain reaction (qRT-PCR) between days 3 and 15. Our secondary objective was to further evaluate how DENV infection history impacts the immunogenicity of the vaccine. Our secondary endpoints were: 1) Change in DENV neutralizing antibody GMT between days 0 and 57; 2) DENV neutralizing antibody GMT at days 0, 28, and 57; and 3) Magnitude of the CD8+ T-cell response at day 15 among groups as measured by activation induced marker (AIM) assays.

### Study design and activities

All details on study procedures are available in the final approved protocol (**Supplementary Data File 2**). For this study, 127 healthy individuals aged 18-59 were screened, 45 were enrolled, 13 were eligible but not needed, and 69 were ineligible (**Extended Data Fig. 1b**). Of those who were ineligible, 39 were DENV naïve but had immunity to other flaviviruses, 13 were unable to comply with the study requirements (typically time commitment), 10 had clinical concerns such as grade 3 bradycardia, hypertension, A1C>8, or taking a biologic medication, and 7 were DENV3 homotypic. At screening, serum samples were collected to assess DENV1-4 immunity by measuring neutralizing antibodies against standard DENV strains using plaque reduction neutralization tests (PRNT). Fifteen individuals were enrolled based on screening PRNT results into each of the three groups: flavivirus naïve, immune to one non-DENV3 serotype (heterotypic), and immune to ≥2 DENV serotypes (polytypic), and this was our intention-to-treat analysis. We pre-specified that our primary analyses would be on neutralizing antibodies measured at day 0, which was felt to be more biologically relevant, as participants were screened up to 6 months prior to vaccination. Additionally, since screening PRNTs were expedited to facilitate enrollment (e.g., conducted as 6-point dilutions), we felt the day 0 PRNTs were more accurate (12-point dilutions, all timepoints run together).

On study day 0, all participants received one dose of the live attenuated rDEN3Δ30/31-7164 with the goal range of 10^3^ ± 10^0.5^ plaque forming units administered as a subcutaneous injection into the deltoid region. Solicited AEs were outlined in the protocol and assessments were performed on days 6, 9, 15, and 28. These days were chosen because they maximized the opportunity to observe a dengue-like rash, and prior studies with this product reported no safety signals prior to day 6 or between days 15 and 28. Unexpected AEs and SAEs were collected through day 180 (required) or day 365 (optional). Blood draws were performed on days 0, 1, 3, 6, 9, 12, 15, 28, 57, 90, 180, and 365 (optional) to evaluate safety laboratory values and trial endpoints.

Participant age, sex, race, and ethnicity were self-reported and recorded by the NIH admissions team upon presentation at the NIH Clinical Center.

### Sample collection

Sera were collected in serum separator (SST) tubes, left vertically for 15-30 minutes on ice, spun at 4°C, 1300g for 10 min, aliquoted into cryovials, flash frozen on dry ice, and stored at -80°C. Sera was heat-inactivated at 56°C for 30 minutes for assays when needed. Peripheral blood mononuclear cells (PBMCs) were collected by drawing whole blood into acid citrate dextrose (ACD) tubes, consolidating it into 50 mL conical centrifuge tubes, and spinning it at 840g for 10 minutes at room temperature. The plasma was removed and stored, and the cell pellet was resuspended with phosphate-buffered saline (PBS) to a final volume of 30 mL. The 30 mL of diluted sample was then layered over 15 mL of Ficoll-Hypaque in a 50 mL centrifuge tube and centrifuged at 1640g for 15 minutes at room temperature. The buffy coat cells were harvested and transferred, washed twice with 45 mL of PBS, and pelleted by centrifuging for 10 minutes at 300g at room temperature. Cells were then resuspended in 90% heat inactivated fetal bovine serum (FBS) + 10% dimethyl sulfoxide (DMSO) and frozen to -90°C using a Thermo controlled rate CryoMed Freezer, Model 7450 at 10 million cells per aliquot. Frozen vials were then transferred for long-term storage in the vapor phase of liquid nitrogen.

### Quantification of vaccine dose

To assure that rDEN3Δ30/31-7164 was dosed at the goal titer of 10^3^ ± 10^0.5^ plaque forming units, leftover product from each administration was analyzed using a vaccine titration assay. The Vero cells (ATCC CCL-81) used for previous titrations of this vaccine in other studies were provided by Dr. Steve Whitehead. An aliquot of the administered vaccine and a positive control virus (DENV4, 1992 Sri Lanka strain, PQ667806) were vortexed and prepared as a 10-fold serial dilution with OptiMEM. 100µL of each diluted virus was then used to infect 90-100% confluent Vero cells on 24 well plates, the plates were incubated for 1 hour at 37°C, and then 1 mL/well of warm overlay media was added (OptiMEM + 2%FBS + 2.5µg/mL amphotericin B + 20µl/mL ciprofloxacin + 1% methylcellulose). Plates were incubated at 37°C for five days. Following this incubation, cells were washed twice with 1 mL PBS then fixed with 0.5 mL of 80% methanol.

Plates were incubated at room temperature for 10 minutes, and frozen at -80°C. To stain for virus plaques, the plates were thawed, methanol removed, 1 mL of antibody diluted in 5% non-fat dried milk buffer in 1x PBS was added to each well, and the plates were incubated for 10 minutes at room temperature. Then, the buffer was removed, 200 µl of a 1:2000 dilution of mouse pan-flavivirus monoclonal antibodies 4G2 and 2H2 antibody was added, and the plates were rocked at 70 rpm and 37°C for 2 hours. The plates were washed twice with 1 mL of antibody dilution buffer, 200 µl of a 1:2000 dilution of secondary horseradish peroxidase (HRP)-labelled goat anti-mouse IgG antibody (KPL/SeraCare, catalog #: 5220-0341, lot #: 10506326) was added, and the plates were rocked at 70 rpm and 37°C for 2 hours. The plates were then washed twice with PBS and air dried for 10 minutes. Then, 180µl of TrueBlue substrate was added to each well, and plates were incubated for 10-15 minutes at room temperature. After incubation, all solution was removed from the plates. The plaques were counted by using a magnifying glass with light immediately or stored at 4°C and counted within 3 days. For each virus sample, the dilution with the best visible plaques were counted. All titrations were performed in duplicate and the average plaque count for each sample was averaged. The virus titer was then calculated by adjusting for the dilution factor of the counted well to obtain the number of plaques per mL. The final titer was reported as the log_10_ of plaques per mL.

Using this quantification method, it was determined that three participants received doses outside of the target range: two received a dose of 10^2.4^ plaque forming units/mL and one received a dose of 10^2^ plaque forming units/mL. Pre-specified sensitivity analyses excluding these individuals from analysis are included in **Supplementary Data File 4**.

### Virus culture measurement for safety evaluation

In the original studies of this product, viral culture was performed to confirm that patient viremia remained at <10^3^ PFU. This assured that there would be minimal risk of severe disease or mosquito transmission of the vaccine product. Given this was the first time that this vaccine product was dosed in individuals with a risk of increased viremia due to antibody-dependent enhancement, we performed a safety run-in where viremia between days 3 and 15 was assessed by viral culture on all individuals until at least two heterotypic participants were vaccinated.

Specifically, sera from participants at days 3, 6, 9, 12, and 15 post vaccination was prepared as a 10-fold serial dilution using OptiMEM. The virus titer from patient sera was then evaluated using the same method described for quantification of vaccine dose. In total, 15 individuals had viremia evaluated by culture, and levels remained at <10^3^ PFU. For trial endpoints, viremia assessments used batched, retrospective measures by qRT-PCR.

In comparing viremia measures by culture to qRT-PCR for the 15 participants tested in both assays, all 9 naïve individuals were positive by qRT-PCR but only 3 by culture at any timepoint. This was expected, given that in previous studies of rDEN3Δ30/31-7164 in naïve participants, 34% experienced viremia by culture, and among those positive, the titer was 10^0.6+/-0.01 48^. Across all individuals who were positive, we found many had culture titers very close to the limit of detection (10^0.5^ to 10^0.7^, depending on the participant), making accurate quantification difficult.

However, qualitatively, viremia positivity measured by qRT-PCR was consistent with culture. For instance, 2/3 heterotypic and 1/3 polytypic were culture positive. For viremia groups defined by AUC viremia and measured by qRT-PCR, 2/3 with high viremia, 1/1 with intermediate viremia, but 0/2 with low viremia were culture positive.

### Adverse events

Solicited, expected, and unexpected AEs were prospectively outlined in the protocol based on prior experience with this product and the typical dengue signs and symptoms, and these included assessments of complete blood counts with differentials, basic metabolic panels, hepatic panels, and coagulation markers, which were performed by the NIH Clinical Center’s Department of Laboratory Medicine according to the schedule of events. AEs were graded per the 2007 FDA guidance: “Toxicity Grading Scale for Healthy Adult and Adolescent Volunteers Enrolled in Preventative Vaccine Clinical Trials.”

Participants experienced mild expected signs and symptoms associated with dengue, blood draws, or typical physiologic variation (such as in heart rate, blood sugar, and potassium). In the first 28 days, there were no grade 4 or unexpected AEs. There were five grade 3 AEs that each occurred one time: hyperglycemia, hypoglycemia, hypokalemia, hemoglobin decrease, and pneumonia. All events were asymptomatic except the pneumonia, which occurred after a known exposure and required oral antibiotics. These grade 3 events were considered not related to the vaccine since these were not observed in prior studies with this product, and dengue is not known to cause these changes. All grade 3 and vaccine related events resolved without sequelae except for the hyperglycemia event, which persisted in a participant who was a known, well-controlled diabetic. There was one severe adverse event, which occurred on day 328 post-vaccination, was unrelated to the vaccine, and consisted of a methicillin resistant staphylococcus aureus (MRSA) infection of the foot after a known exposure that required hospitalization for intravenous antibiotics and debridement. This infection was resolved without sequelae at the last participant contact.

### Additional biomarkers

The protocol was amended to include the evaluation of C-reactive protein (CRP), ferritin, immunoglobulins (IgG, IgA, IgM), fibrinogen, IgE, and tryptase by the NIH Clinical Center’s Department of Laboratory Medicine. Because these biomarkers have been identified as altered in severe dengue ^49, 50, 51, 52, 53^, we assessed whether they varied after vaccination. Once the amendment was approved, these biomarkers were collected on 31 participants.

### qRT-PCR

Genomic RNA was extracted from serum samples using EZ1 Advanced XL System (Qiagen) and reagents from Qiagen viral RNA mini kit (EZ1 Virus Mini Kit v2.0, 955134) following the manufacturer’s protocol. Complementary DNA synthesis was performed by reverse transcription using 4μL of superscript IV Vilo Master Mix (11756050) in 16μL extracted RNA for a final reaction volume of 20μL per sample. Samples were run on the C1000 Touch™ Thermal Cycler (Bio-Rad Laboratories, 1851197) with the following reaction conditions: 25°C for 10 minutes, 50°C for 10 minutes, 85°C for 5 minutes and held at 4°C. The following reagents were used for master mix preparation: reverse transcriptase (TaqMan Fast Universal PCR Master Mix [2X], no AmpErase UNG, Invitrogen 352042), The NS2A region of the DENV genome was amplified using consensus primers D3-3620F (5’TCCTTCTCTCAGGGCAAATAAC-3’,100 mM, Integrated DNA Technologies), D3-3701R (5’ CTAGGTAAGTGACGCCCATTC -3’, 100 mM, Integrated DNA Technologies), D3-3669P (5’AATGATTGGGTCCAACGCCTCTGA -3’, TaqMan fluorogenic probe 100 mM, Dye: 5’ 6-FAM™ Quencher: 3’ Black Hole Quencher®1, Integrated DNA Technologies) and nuclease-Free Water-not DEPC-Treated (Life Technologies AM9937). Eight ten-fold serial dilutions starting 1:10 of standard curve of cDNA plasmid (pDEN3D30/31 for DENV3, 2ng/µL) was prepared in triplicate in each experiment. All samples were prepared in MicroAmp™ Splash-Free 96-Well Base (Life Technologies 4312063) and quantified using QuantStudio 6 Flex Instrument (Applied Biosystems) under the following amplification conditions: initial denaturation at 95°C for 20 seconds, followed by 40 cycles of melting 95°C for 1 second, annealing and extension 60°C for 20 seconds. Samples were tested at days 3, 6, 9, 12, and 15. To estimate the viral titer (Copy Number per mL), we used the following equations. Firstly, Copy Number for first dilution of the standard was calculated as follows and we divided by 10 for subsequent dilutions,

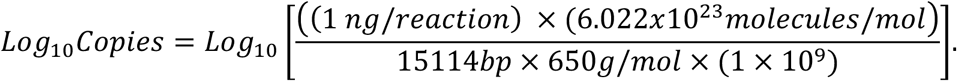

Linear regression of the Log10 Copy Numbers (CN) versus the CT values for the standard was generated. The slope and y intercept of the resulting plot was used to calculate the Copy Number per reaction for samples (CN/reaction).

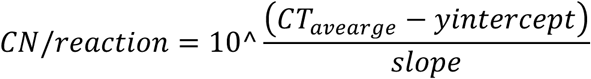

We then used the following equation to calculate the Copy Number per mL (CN/mL) for samples,

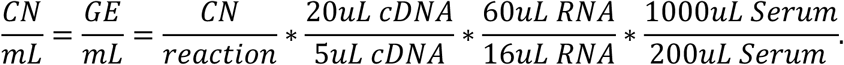

To estimate viral burden, we calculated the area under the curve (AUC) for the viral titer (genome equivalent/mL) across study days 3 to 15.

### Standard and mature DENV1-4 plaque reduction neutralization test (PRNT)

Low passage clinical DENV isolates were used in all experiments, as previously described (GenBank accession numbers PQ667803, PQ667804, PQ667805, and PQ667806, viruses originally provided by Dr. Aravinda de Silva). Viruses were prepared using ‘standard’ conditions by amplifying on Vero cells (ATCC CCL-81) provided by Dr. Eva Harris of the University of California, Berkeley. To prepare mature forms of the same viruses, each DENV isolate was amplified on Vero cells (ATCC CCL-81) engineered to over-express furin ^54, 55^ provided by Dr. Ralph Baric of University of North Carolina Chapel Hill. Vero cells were maintained at 37°C and 5% CO_2_ with 90% humidity in Opti-Pro serum-free media (Invitrogen) supplemented with 10% FBS and 4 mM glutamine. The neutralization assay was conducted as previously described^27^.

Vero cells were plated at 1.7x10^4^ per well allowed to expand and form a monolayer at 37°C overnight. Sera were serially diluted in six four-fold dilutions (1:10 to 20480) for screening neutralization assays or twelve two-fold dilutions (1:10 to 1:20480) for trial endpoint assays as technical duplicates, mixed with virus, and incubated at 37°C for one hour. The serum-virus mixture was then used to inoculate confluent Vero cells for one hour at 37°C. Thereafter, 1% methycellulose overlay media was added, and plates were incubated for two days to allow plaque formation. Cells were washed with 1X PBS, fixed with 80% methanol, blocked with 5% non-fat dried milk (10 minutes), and immunostained using mouse pan-flavivirus monoclonal antibodies 4G2 and 2H2 each diluted 1:2000 in 5% non-fat dried milk for two hours at 37°C. Thereafter, cells were washed, and secondary HRP-labelled goat anti-mouse IgG antibody diluted 1:3000 (KPL/SeraCare, catalog #: 5220-0341, lot #: 10506326) was added to cells and incubated for one hour at 37°C. Plates were washed with 1X PBS, allowed to dry, and developed using TrueBlue HRP substrate until plaques resolved. Images of wells were collected using the Cellular Technology Limited (CTL) machine and ImmunoSpot software. Automated plaque counting was conducted using the Viridot plaque counter package in R^56^. Plaque count data were normalized relative to the virus only control and fitted with logistic regression using the drc package in R to estimate the PRNT_50_ titer, the point at which a 50% reduction in plaque count was observed relative to the virus only control wells. A serum from a participant with broadly neutralizing responses and a history of dengue was used as a positive control, and serum from a DENV-negative participant as a negative control. GMTs were calculated as the average of the log transformed DENV1-4 neutralizing antibody titers. Samples were tested at screening (standard neutralization assay) and days 0, 28, and 57 for standard and mature neutralization assays.

Breadth of neutralization was defined as number of DENV serotypes with neutralization titers ≥1:60 for either the standard or mature neutralization assay.

### Flavivirus PRNTs

To evaluate neutralization to flaviviruses, the PRNT was conducted as described above for standard virus. The following viruses were provided by Dr. Steve Whitehead: ZIKV-Paraiba/2015, (GenBank accession number: AF326573); WNV/DEN4Δ30, which contains E and prM proteins of WNV (AF196835) within a DENV4 backbone (AF326827), JEV vaccine strain SA14-14-2 (MH258849), and the yellow fever 17D vaccine strain (X03700.1). For screening, sera were tested only at a 1:10 and 1:40 dilution for those who were negative by DENV1-4 neutralization assay. Day 0 neutralization titers were tested for all DENV-immune participants in 12-fold serial dilutions (1:10 to 10:20,480). Given differences in plaque size, ZIKV and JEV were fixed after 40 hours and YFV and WNV at 48 hours. A human serum negative to all flaviviruses and a positive control specific to each flavivirus was included in each experiment.

All staining was conducted as described above, except that because JEV is not efficiently stained by 4G2 and 2H2, an in-house JEV-specific mouse serum diluted 1:3000 was used as the primary antibody.

### Peptide synthesis and megapool preparation

Peptide megapools were designed and synthesized to allow simultaneous testing of numerous T cell epitopes, as previously established^57, 58^. Briefly, individual peptides were synthesized as crude material (TC Peptide Lab, San Diego, CA) and solubilized in DMSO at a concentration of 10-20 mg/mL. Peptides were pooled into their corresponding megapools, sequentially lyophilized to reduce cytotoxicity and resuspended in DMSO at a final concentration of 1 mg/mL. The DENV megapools used in this study were derived from predicted and experimentally defined DENV epitope pools from the four DENV serotypes, namely, a CD4^+^ T cell megapool comprising 180 fifteen-mers^59^ and a CD8^+^ T cell megapool comprising 268 nine-and ten-mers ^60^. The total CD4^+^ and CD8^+^ DENV T cell epitopes were divided into three different megapools to discriminate responses by viral region. One structural megapool was composed of epitopes targeting structural DENV proteins (C, prM, E), and two non-structural megapools encompassed the epitopes NS1, NS2A, NS2B, and NS3 and separately, NS4A, NS4B and NS5.

### Activation-induced markers (AIM) and intracellular cytokine staining assay

PBMCs were stimulated ex-vivo to enable the examination of antigen-specific T cell responses using flow cytometry. First, cryopreserved PBMCs were thawed by diluting in 10 mL pre-warmed complete RPMI 1640 with 5% human AB serum (Gemini Bioproducts) and 50 U/mL benzonase (Sigma-Aldrich, St. Louis, MO) and centrifuged at 1200 rpm for 7 minutes. Each participant’s sample was prepared in five different conditions, each at 1x10^6^ PBMC per well in 96-well U bottom plates in complete RPMI containing 5% Human AB Serum. Cells from each sample were stimulated for 24 hours under the five conditions: the three DENV megapools at 1 µg/mL, phytohaemagglutinin (PHA) at 1 µg/mL as positive control, and an equimolar amount of DMSO as a negative control. Fifteen minutes prior to the addition of stimuli, cells were treated with 5 µg/mL anti-human CD40 antibody. The complete list of antibodies and reagents used in this panel can be found in **Supplementary Fig. 1**.

During the last 4 hours of stimulation, cells were treated with Golgi-Plug and Golgi-Stop, along with anti-human CD69 and CD137 antibodies. Following stimulation, supernatants were collected, cells were washed, incubated with human Fc-block, and stained with LIVE/DEAD viability marker for 15 minutes according to the manufacturer’s instruction (Thermo Fisher).

Subsequently, cells were washed and stained with anti-human antibodies (CD3, CD8, CD4, CD14, CD16, CD20, CCR7, OX40, CD45RA) in PBS with 3% FBS and 10% Brilliant Staining Buffer Plus for 30 minutes at 4°C. Finally, cells were fixed in 4% paraformaldehyde (PFA) (Sigma-Aldrich) for 10 minutes at 4°C, followed by washing with permeabilization buffer (PBS with 5 µg/mL saponin, 10% bovine serum albumin and 1% azide) at room temperature. For intracellular staining, cells were stained with anti-human antibodies (IFN-γ and CD40 Ligand) in permeabilization buffer with 10% human AB serum and Brilliant Staining Buffer Plus for 30 minutes at room temperature. Samples were acquired on a Cytek Aurora flow cytometer (Cytek Biosciences, Fremont, CA). Samples were analyzed using FlowJo 10.10 software. A representative gating strategy is shown in **Supplementary Fig. 1**. AIM positive-specific T cells are defined for co-expressing the markers OX40^+^CD137^+^ for CD4^+^ T cells and CD69^+^CD137^+^ for CD8^+^ T cells as previously described^10^. IFN-γ-producing antigen-specific T cells were defined by their production of IFN-γ along with the co-expression of CD40L for CD4^+^ and CD69 for CD8^+^ T cells. T cells were grouped by memory subset as follows: Naïve (Tn) as CD45RA^+^CCR7^+^; central memory (Tcm) as CD45RA^-^CCR7^+^; effector memory (Tem) as CD45RA^-^CCR7^-^, and effector memory re-expressing CD45RA (Temra) as CD45RA^+^CCR7^-^.

Data were normalized with a minimal level of sensitivity of 0.005, and the percentage of virus-specific T cell responses is shown as background (DMSO) subtracted data. A limit of detection (LOD) and limit of sensitivity (LOS) were calculated for each population. LOD was calculated as the upper 95% confidence interval of the negative control (DMSO) values. LOS was calculated as median + 2x Standard Deviation (SD) of negative control (DMSO) values.

Stimulation Index (SI) was also calculated for each sample and population, as % of response divided by % of response in DMSO control. Responses > than LOS and SI > 2 were considered positive. Responses with SI < 2 were normalized to LOD. Samples with low viability (under 60%) or poor activation response to PHA stimulation were discarded: this included one day 0 and one day 15 sample. One other sample could not be run at day 28 due to low PBMC volume. Percentages were summed across peptide pools to estimate total DENV-reactive T cells for each participant. Each memory T-cell subset, originally reported as a proportion of the total CD4+ and CD8+ AIM+ population, was multiplied by the CD4 or CD8 AIM+ samples to get the magnitude of the memory subsets. Values at the LOD were set to 0 and, for certain analyses, non-structural and structural DENV antigens were added to get a single value for each memory subset. Other analyses included comparisons of non-structural and structural antigens and, in this case, only the non-structural antigens were added. Values at 0 were then set to the half of LOD and all data was log_10_ transformed.

### Antibody-dependent enhancement assays

Day 0 participants serum and a separate DENV-seropositive (DENV2 monotypic) plasma sample (positive control) were assessed for ADE activity using U937 cells expressing CD16^61^ and K562 cells. U937+CD16 cells were provided by Dr. Stylianos Bournazos of Rockefeller University and K562 cells were provided by Dr. Theodore Pierson of the NIH. U937+CD16 cells were passaged three times a week and maintained in complete RMPI media (RPMI 1640 media supplemented with 10% FBS, 2 mM of L-glutamine, 100 U/mL penicillin, 100 ug/mL streptomycin, 5 ug/ml of Blasticidin (Gibco)) at 37°C and 5% CO_2_ with 90% humidity.

Blasticidin was supplement fresh each passage. K562 cells were passaged three times a week and maintained in RPMI 1640 media supplemented with 7% FBS, 2mM of L-glutamine, 100U/mL penicillin, and 100ug/mL streptomycin.

Heat-inactivated sera were serially diluted in complete RPMI media (eight four-fold dilutions starting at 1:10). 50uL of diluted serum was pre-incubated with DENV3 virus (rDEN3Δ30/31-7164) (5×10^3^ infectious units/well corresponding to MOI=0.1) at 37°C for 1 hour in 96-round bottom plates. 5x10^5^ cells/mL U937+CD16 and K562 cells were prepared and resuspended in complete RPMI medium. 100uL of the 5x10^5^ cells/mL was added to a total volume of 200uL for a final concentration of 2.5x10^5^ cells/mL (50uL sera, 50uL virus, 100uL cells). Additional, controls included virus only (RPMI with virus without serum sample; six four-fold dilutions starting at 1:10) and cell only conditions (RPMI only without virus and without serum sample; in replicate). All wells were prepared in duplicate. Plates were then incubated at 37°C for 48 hours. Two days post-infection, supernatant was carefully removed from the cell pellet. Cells were washed twice with PBS containing 1% FBS and centrifuged at 500g for 5 min at 4°C. After centrifugation, supernatant was carefully removed from the cell pellet and then fixed and permeabilized using the Cytofix/Cytoperm Fixation/Permeabilization Kit (BD Biosciences). Cells were fixed using 100ul of Cytofix/Cytoperm buffer and incubated for 30 min at 4°C. After permeabilization, the cells were washed twice with the permeabilization buffer (1x PermWash) and centrifuged at 500g for 5 minutes at 4°C. Supernatants were carefully removed from the cell pellet and 100 ul of a custom AlexaFluor647-conjugated mouse anti-dengue 4G2 (0.25 ug/ml) diluted in permeabilization buffer was added to each well and incubated for 60 minutes at 4°C. Each well was then washed twice with 150 ul of permeabilization buffer (1x PermWash) and centrifuged at 500g for 5 min at 4°C. Supernatant was removed and cells fixed with 200ul of 1% paraformaldehyde (Electron Microscope Sciences) diluted in PBS containing 1% FBS. Samples were stored at 4°C and later analyzed using a Guava easyCyte flow cytometer (Cytek). Data were analyzed using FlowJo 10.10.0 software. A representative gating strategy is shown in **Supplementary Fig. 2**. Fold-enhancement was calculated by dividing percent infection at each serum dilution by the mean of the mock controls on each plate. Replicates on each plate were averaged. Peak enhancement titers were defined as the dilution that produced maximum fold-enhancement. One participant had missing K562 enhancement measures.

### E dimer epitope (EDE) blockade of binding ELISA

We used a blockade of binding ELISA assay to measure human antibodies that block binding of monoclonal antibody EDE1 C8^11^ to stabilized forms of the envelope dimer proteins. The envelope dimers were purchased from UNC Protein Expression and Purification Core from constructs provided by Dr. Aravinda de Silva. The assay was performed as previously described^8^. High-affinity 96-well flat-bottom microtiter plates (Microlon, Greiner, Germany) were coated with 100ng of anti-His monoclonal antibody (TA150088, OriGene) in 1x Tris-buffered saline (TBS: 50 mM Tris-Cl at pH 7.5 containing 150mM NaCl) and incubated overnight at 4°C. Plates were washed 3x using TBS-T wash buffer (1x TBS with 0.05% Tween 20 [P7949, Sigma Aldrich]), blocked using TBS-T containing 5% Carnation Instant Non-fat Dry Milk (12428935), and incubated for 1 hour at 37°C. Plates were washed and incubated with 100 ng of stabilized E dimer antigens in 5% milk TBS-T, produced as previously described, and incubated for one hour at 37°C. Each antigen was tested separately: wild-type World Health Organization reference strain sequences for DENV1 (SC.12), DENV2 (SC.14), DENV3 (SC.12), or DENV2 with a mutation in the fusion loop at position 106 (SC.10)^62, 63^. All antigens had been previously avi-biotin labeled for other purposes. After wash step, 50 μl of serially diluted human plasma samples (1:10, 1:100, 1:1000, and 1:10,000 dilutions in blocking buffer, in duplicate), and positive (DENV-exposed human donor) and negative (DENV-naïve human donor) controls were added and plates incubated for two hours at 37°C. Plates were washed and alkaline phosphatase (AP)-conjugated DENV EDE1 C8 human monoclonal antibody (TP41002F) was added to each well at a final concentration of 2000 ng/ml (100 ng total). Labeled antibody was prepared in-house using an Alkaline Phosphatase Conjugation Kit-Lightning-Link (ab102850, Abcam) following manufacturer guidelines, and was incubated with the samples for one hour at 37°C. Following another washing step with TBS-T, 50 μl of alkaline phosphatase substrate (SIGMAFAST, N2770-50SET, Sigma-Aldrich) was added to each well and color was allowed to development for 30 minutes at room temperature in darkness. The enzymatic reaction was terminated by adding 50 μl of 1N sodium hydroxide solution (SS255-1, Fisher Chemical) to each well, after which the optical density (OD) values were measured at an absorbance of 405 nm using a Cytation 7 microplate reader. We calculated binding inhibition activity as the percentage reduction in EDE1 C8 binding by comparing test samples to dengue-naïve human serum (negative control) using the formula: Percent inhibition = 100 – ((OD of sample / OD of negative control) × 100). EDE antibody titers were estimated by 4-parameter logistic regression analysis in RStudio to calculate the IC50 values (the dilution causing 50% reduction in EDE1 C8 binding). For each participant, titrations against all antigens were performed within the same experimental run for all timepoints (day 0, 28, and 57) to minimize variability.

### IgG antibodies to DENV1-4 EDIII and NS1 proteins

IgG antibodies to the envelope protein domain 3 (ED3) of DENV1-4 and ZIKV and nonstructural 1 proteins (NS1) of DENV1-4, ZIKV and YFV were measured on day 0 serum samples from all participants using a Luminex multiplex assay as previously described^27, 64^. MagPlex®-Avidin Microspheres (Luminex) were coated with biotinylated ED3 antigens or biotinylated bovine serum albumin (BSA), while avidin-coated microspheres were coated with anti-His tag antibody (abcam) to capture his-tagged NS1 antigens (The Native Antigen Company). Antigen-conjugated microspheres were mixed and a final count of 2,500 beads per antigen were added to each well. Human sera were diluted 1:200 and 1:500. The dilution 1:500 has been validated previously^64^ and was used for our final definitions, and 1:200 was run as a sensitivity analysis because our population includes those with more remote prior infections.

Diluted serum was incubated in singlicate with the microspheres for one hour at 37°C and spun at 700rpm. Serum-bead mixtures were washed 3x then incubated with 50 µL goat anti-human IgG Fc multi-species SP ads-PE antibody (Southern Biotech, catalog #: 2014-09). Antibody binding was measured on a Luminex 200 analyzer and estimated as median fluorescence intensity (MFI). Positivity was estimated for each antigen separately and defined the mean + 3 standard deviations for 130 negative control sera tested at a 1:500 dilution. Data were analyzed directly as MFIs and as number of DENV1-4 ED3 or NS1 antigens to which an individual had a positive response.

### Travel history and exposure surveys

Participants completed surveys in REDCAP prior to screening and following enrollment to evaluate prior flavivirus exposure and vaccination history. Participants provided information on when how long they had lived in or traveled to countries defined as having “frequent or continuous” or “sporadic or uncertain” dengue risk based on maps provided by the Centers for Disease Control (https://www.cdc.gov/dengue/areas-with-risk/index.html). They also provided history of vaccination with yellow fever, Japanese encephalitis, and tick-borne encephalitis, as well as any experimental vaccines for other flavivirus vaccines, which were an exclusion rule. Detailed information on prior dengue, Zika, other flavivirus, and febrile illnesses were surveyed, along with information on known contacts exposed to dengue, and individual and community efforts to control dengue in areas where they had lived or traveled. The following variables were constructed from these surveys. Time since having a dengue case or the last time a participant lived in an endemic area was quantified relative to their vaccination date. High dengue risk was defined as knowing one or more people who lived in the same region (100 miles) or having a family member living in the same house who got dengue. Those with community risk were those who said their community took steps to control dengue by managing standing water, applying pesticides, or through public awareness campaigns. Any prior flavivirus vaccine or exposure was defined as any of the following: evidence of prior yellow fever vaccination (PRNT > 1:10 or prior vaccine), Zika (self-reported case, or PRNT titer >1:100 and ZIKV NS1 positive at 1:500 dilution), Japanese encephalitis vaccine (PRNT >1:10 or prior vaccine), and history of tick-borne encephalitis vaccine. There were no flavivirus infections reported other than Zika.

### DENV3 E dimer IgG ELISA

IgG binding antibodies to the DENV3 stabilized E dimer were measured in plasma from all participants at days 0, 1, 3, 6, 9, 12, 15, 28, and 57 and was performed as described above for the EDE blockade of binding assay, with the following differences. Following incubation with serum, wells were stained with AP-conjugated anti-human IgG (1:2000 dilution; Sigma-Aldrich) for 1 hour at 37°C. As above, plates were washed and developed with AP substrate and OD was measured after leaving plates for 15 minutes. ELISA signals are reported as the OD of the average of duplicates after subtraction of the negative control sample. DENV naïve human serum served as a negative control, and DENV monoclonal antibody C8 was used as a positive control. For the E dimer ELISA, serum titers for 50% binding activity (effective concentration 50, EC_50_) were calculated by nonlinear, dose-response regression using the drc package in R.

### Cytokine analysis

Cytokine and chemokines abundance in participants’ serum were assessed using Luminex Human Cytokine Magnetic 30-Plex Panel for Luminex^TM^ Platform according to manufacturer’s instructions. MAGPIX^TM^ instrument was used for data acquisition. Briefly, for each analyte, a nine-point dilution series of the protein standard plus an assay diluent-only background were run in duplicate. Supernatant (positive control) from Phorbol Myristate Acetate and ionomycin (PMAi)- stimulated PBMCs and plasma from a healthy donor (negative control) were run in duplicate. Patient samples (n=40 participants) were run in duplicate for study days 0,1,3,6,9,12,15 and 28. Invitrogen ProcartaPlex Analyst software version 1.0 was used for data analysis where a five-parameter logistic model was used for standard curves and to quantify protein concentrations.

### ELISpot for measuring plasmablasts

ELISpot assay was run on fresh PBMC samples. With the aim of running two participants per vaccination block with different infection histories, plasmablast responses were only assessed in a subset of individuals that included n=5 naive, n=7 heterotypic, and n=7 polytypic individuals. Human IgM Single-Color ELISPOT and Human IgA/IgG Double-Color ELISPOT kits, which include 96-well plates, capture, detection, tertiary and substrate solutions and diluents, were used for plasmablast detection (IMMUNOSPOT by C.T.L). Total Ig or DENV-reactive antibody secreting cells were detected using a combination of anti-human IgK and IgL (IgK/L) or DENV3 or DENV1,2,4 E protein antigens. Given timing of when the assay was started and lack of availability of the stabilized E dimers, wild-type E proteins were obtained commercially (Native Antigen Company) and included DENV1 (FGA/NA strain, REC31679), DENV2 (New Guinea C strain, REC31680), DENV3 (H87 strain, REC31681), and DENV4 (Philippines/H241 strain, REC31682). Antigen and IgK/L (250 ng/well) was prepared using Diluent A reagent provided by the manufacturers. 80uL of the antigen and IgK/L capture solutions were added to sample wells and IgK/L was also added to both healthy donor and blank wells, which were used as controls in each experiment, and incubated overnight (12 hours) in 4°C. The coating solution was decanted and 150 uL/well of sterile PBS was added to remove excess unbound antibody or antigen and later decanted. Assay media was prepared (RPMI supplemented with 10 % FBS, 2mM of L-glutamine, 100U/mL penicillin, 100ug/mL streptomycin, 0.1g of HEPES, and 0.05mM of 2-mercaptoethanol) and 150 uL was added to each well and plates were incubated for one hour at room temperature. Isolated fresh PBMCs from participants (days 9 and 15) and cryopreserved PBMCs of healthy donor were plated at 5x10^5^, 2.5x10^5^ and 1.25x10^5^ cells/well for antigen-reactive antibody-secreting cell detection and 1x10^5^ and 5x10^4^ cells/well for total Ig antibody-secreting cell detection. 200uL of PBMCs suspension was added to each well with cells and 200 uL of assay media was added to each blank well and incubated at 37°C for four hours. Plates were then washed twice with PBS (pH 7.2) followed by PBS containing 0.05% Tween-20 (PBS-T). The solution was removed, 80uL of anti-human IgM and IgG/IgA detection solution was prepared per manufacturer’s protocol and added to respective wells, and plates were incubated in the dark at room temperature for two hours. 80uL of tertiary solution was added per well in both IgM and IgG/IgA ELISpot plates. The IgM plate was incubated in the dark at room temperature for 30 minutes and thereafter plate was decanted and each well washed twice with 150 uL of distilled water. 80 uL of CTL-TrueBlue Substrate Solution was added to each well and incubated in the dark at room temperature for 10 minutes. The solution was removed; plates were washed twice and then dried in the flow hood. The IgG/IgA plate was incubated in the dark at room temperature for one hour and thereafter the plate was decanted and each well washed twice with 150 uL of distilled water. 80 uL of CTL-TrueBlue Substrate Solution was added to each well and incubated in the dark at room temperature for 10 minutes. Plates were decanted and each well washed twice with 150 uL of distilled water. 80 uL of CTL-TruRed Substrate Solution was added to each well and incubated in the dark at RT for seven minutes. Plates were decanted, washed twice with 150 uL of distilled water, and dried before imaging and spot counting using the CTL Immunospot software. Of the 19 participants run, one participant did not have a day 9 timepoint.

### Statistical analyses

All statistical analyses were conducted in R 4.3.3 (R Foundation for Statistical Computing). Statistical analyses were planned as described in **Supplementary Data File 3**. All participants who receive the single dose of vaccine were analyzed for safety and immunogenicity. All continuous biological data were log_10_ transformed unless otherwise stated. Observations below the assay lower limit of detection (LLOD) were set to a value of half the LLOD. Continuous data are presented as means with standard deviations, or model coefficient estimates with confidence intervals. All participants completed the study, but a few missed a visit. For variables with missed visits when not performing modeling (e.g., summary measures like peak viremia), these data were considered missing completely at random and estimates used available data. When modeling repeat measures or multiple predictors, data were also considered missing at random and imputed as the median value for that variable to enable inclusion of the participant in the larger model.

For primary endpoints, we used Firth’s penalized logistic regression to estimate the odds of experiencing each AE, evaluating the overall model p-value using the likelihood ratio. For those significant at p<0.05, then we evaluated the coefficients and p-values for immune group comparisons from the same model. Safety tests were performed at a 0.05 level of significance, except rash, was performed at a 0.05/3 level of significance. The fold change in the DENV neutralizing antibody GMT from day 0 to 28 in each group was planned to be assessed using a paired t-test with a 2-sided 0.05 level test, but due to lack of normality after testing by Shapiro-Wilk test, an unadjusted Wilcox signed-rank test was used. Mean peak viremia from days 3 and 15 among groups was assessed by t-tests with Bonferroni correction, after confirming normality.

For secondary endpoints, the fold-change in the DENV neutralizing antibody GMT from day 0 to 57 in each group was also tested using Wilcox signed-rank tests with a 2-sided 0.05 level test. The magnitude of activated CD8^+^ T cells at day 15 was compared between groups with each test performed at the 2-sided 0.05 level. Linear and non-linear regression models of day 28 and 57 DENV neutralizing antibody GMTs were also performed to evaluate differences by day 0 DENV neutralizing antibody GMT, adjusting for demographic variables and time since dengue exposure. Results for intention to treat primary and secondary analyses are described in **Supplementary Data File 4.**

We pre-specified that we would evaluate clustering among variables that define dengue-like illness or viremia to identify associations with baseline and acute-phase immunity regardless of infection history group. While our primary endpoints focused on peak viremia, we felt viremia burden measured as AUC of viremia better captured the entirety of the response to the vaccine. Partitioning around medoids (PAM) was used to cluster AUC viremia to identify differential vaccine responses among DENV-immune participants. To determine the optimal number of clusters, silhouette plots were inspected for solutions ranging from k=2 to k=4. The three solutions yielded relatively similar average silhouette widths that is, S=0.64, 0.63 and 0.65 respectively. However, a visual inspection of the plots confirmed that k=3 provided well-defined structure, with no negative silhouette scores and most points exhibiting high positive scores.

All exploratory immunological analyses were considered exploratory and performed at a 2-sided 0.05 level of significance, as planned. We initially planned to use parametric tests, but following examination of the data and determining the data often had skewed distributions, non-parametric tests were used. Kruskal-Wallis was used for comparisons across groups, and if significant at p<0.05, followed by Dunn’s test, with Bonferroni corrections when multiple comparisons were conducted. Differences between paired measures were evaluated with Wilcoxon-signed rank test. Wilcoxon rank-sum test was used for each immune group comparison to the naïve and Bonferroni correction was not conducted as the naïve group was considered the reference group. Spearman’s correlation coefficients were used to estimate correlations between variables.

Regression was used to adjust for demographic variables (age, sex, race, and ethnicity), and continuous predictors were evaluated both for linear and non-linear associations with endpoints using generalized linear models assuming gaussian error distributions and generalized additive models. For measures of general biological responses and not DENV-specific responses, like biomarkers and cytokines, we performed analyses after subtracting day 0 values for each participant to enable estimation of change due to the vaccine.

The immune correlates of viremia analyses were conducted as follows. Immune measures and exposure history variables (“measures”) collected at baseline were evaluated as predictors of AUC viremia and viremia groups (naïve, high, intermediate, and low viremia). Measures included: infection history groups, number of contacts with dengue, community dengue risk, having a prior non-DENV flavivirus exposure or vaccine, time since having dengue, time since living in a dengue-endemic area, and total time living in an endemic area. Neutralizing antibodies were measured separately against (1) mature DENV1-4, (2) standard DENV1-4, and (3) standard JEV, WNV, YFV, and ZIKV. The following metrics were estimated for each of these three virus groupings: titer to each virus and GMT across viruses; breadth (number of serotypes) neutralized at a 1:60 dilution; and virus with the maximum observed titer. Binding to ED3 and NS1 of DENV1-4 (and ZIKV and YFV for NS1) by serum at a 1:500 dilution was measured as MFI, and breadth was evaluated as number of viruses bound for each protein above the threshold of positivity. Antibodies that competed with the EDE antibody for binding to stabilized dimers of DENV1, DENV2, DENV3, and DENV2 with mutation in the fusion loop were measured as titers. Enhancement of DENV3 on U937 + CD16 and K562 cells was quantified as enhancement at a 1:10 dilution and peak enhancement titer. IgG binding antibody IC_50_ values were measured to the DENV3 dimer. Magnitude of AIM^+^ T cells was measured separately for CD4^+^ and CD8^+^ cells, and quantified as total cells, and by memory subset (naïve, central memory, effector memory, and Temra) and IFN-γ^+^ cells. The sum of all T cells was also included as a predictor for total cells, TEMRA, and IFN-γ^+^ cells. Missing data included one individual without T cell data at baseline, and one with missing K562 enhancement titers. Values were set to the median value for that immune measure.

We first evaluated the linear and non-linear association of each measure with AUC viremia. All measures were tested as univariable predictors and in models adjusted for demographic variables (age, sex, race, and ethnicity). Only adjusted model results are shown. Linear associations were evaluated using a generalized linear model assuming a gaussian error distribution. We report effect sizes and confidence intervals for all predictors with their associated p-values using forest plots (**Fig. 3a**, **Extended Data Fig. 6**). Non-linear associations for each predictor were estimated using the generalized additive models in the mgcv package in R, using the s() smooth and basis dimension (k) set to 4 to enable us to capture strong non-linear effects. Both linear and non-linear associations are visualized in **Extended Data Fig. 7**.

We also evaluated the set of measures with the strongest ability to predict AUC viremia and viremia groups. Data were standardized and scaled prior to analysis to enable comparison of effects across measures. To identify variables with linear associations with viremia, we used elastic net regularization, which balances LASSO and ridge regression to enable variable reduction while accounting for collinearity of variables. Elastic net was performed using glmnet in the train function from the caret package. A generalized linear model was used to predict AUC viremia, and multinomial regression to classify viremia groups. Random forest models were used to identify the most important variables even with non-linear associations to AUC viremia and viremia groups. Both the elastic net and random forest models were trained using random repeat cross-validation with 10 repeats each of size 10, and a random search. The tunelength was set to 100. Model comparison was performed using the resamples function in caret. The elastic net model performed slightly better for predicting AUC viremia, while random forest performed slightly better for viremia group classification. Given the simpler interpretability of the elastic net model, we focus on those results but present the random forest models to identify immune measures with non-linear associations that may modestly improve prediction.

**Extended Data Fig. 1.**
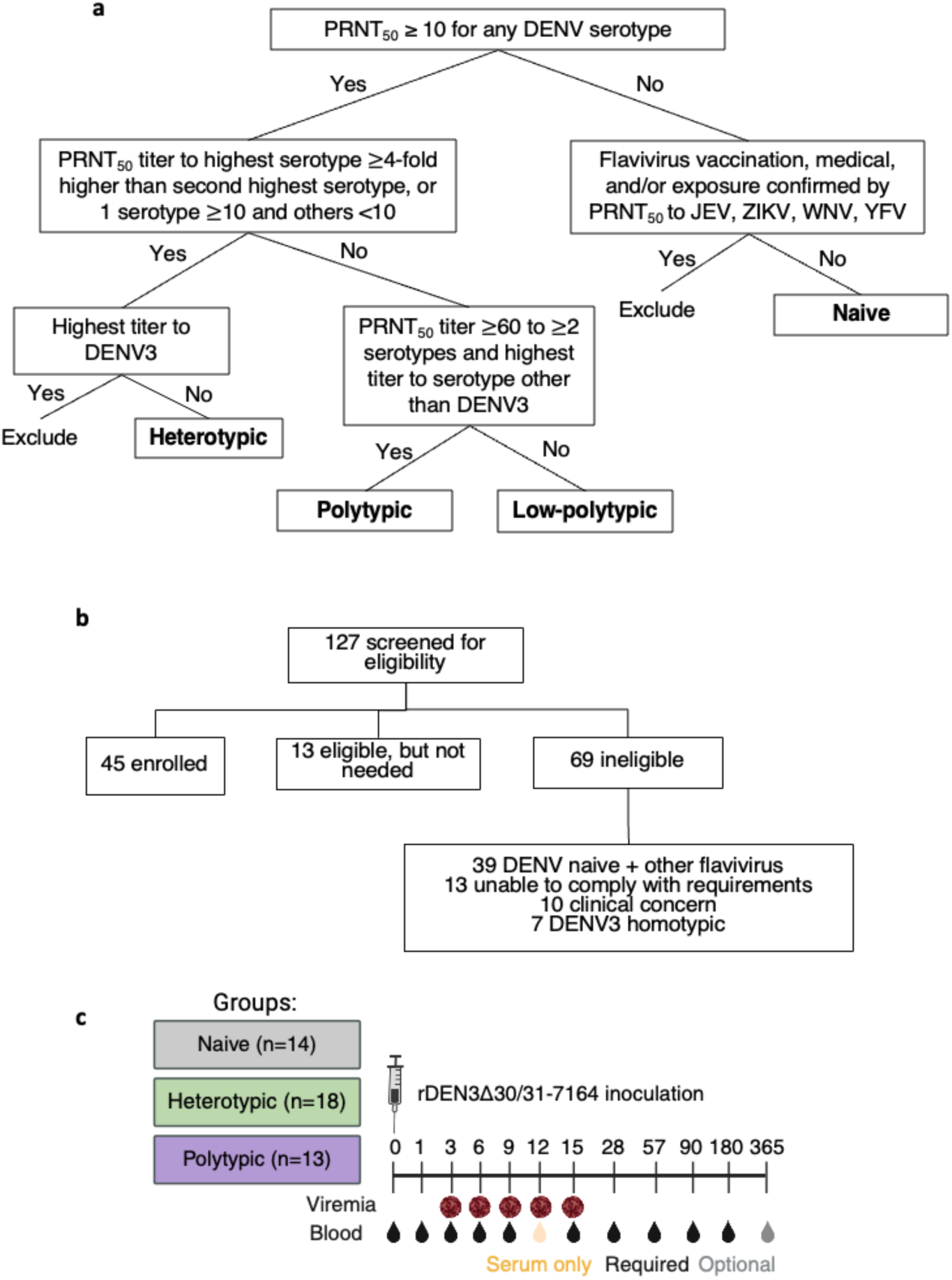
Serological criteria, participant screening, and study design. **(a)** To define serology groups, all participants were tested by PRNT for neutralizing antibodies to standard preparations of DENV1-4, which are only partially mature. The PRNT_50_ is the titer at which 50% reduction in plaque count was observed relative to a negative control serum. Only DENV1-4 PRNT_50_ negative participants were tested by neutralization assay to flaviviruses, including Japanese encephalitis virus (JEV), Zika virus (ZIKV), West Nile virus (WNV), and yellow fever virus (YFV). The criteria in this flow chart were applied to screening neutralizing antibody testing, which was conducted between zero and six months before vaccination, and day 0 neutralizing antibody testing, conducted on the sample collected on the day of vaccination. In addition to including those with low balanced titers, the low-polytypic group was allowed to include those with the highest titer to DENV3 but no individuals meeting these criteria were enrolled. **(b)** A total of n=127 individuals were screened for clinical appropriateness and neutralizing antibodies. Clinical concerns resulting in exclusion included grade 2-3 abnormalities in heart rate, blood sugar, liver tests, hemoglobin, hypertension, autoimmune diseases, and clinically significant acid reflux. **(c)** Final enrolled groups per day 0 antibody titers are depicted with sample collection timepoints.

**Extended Data Fig. 2.**
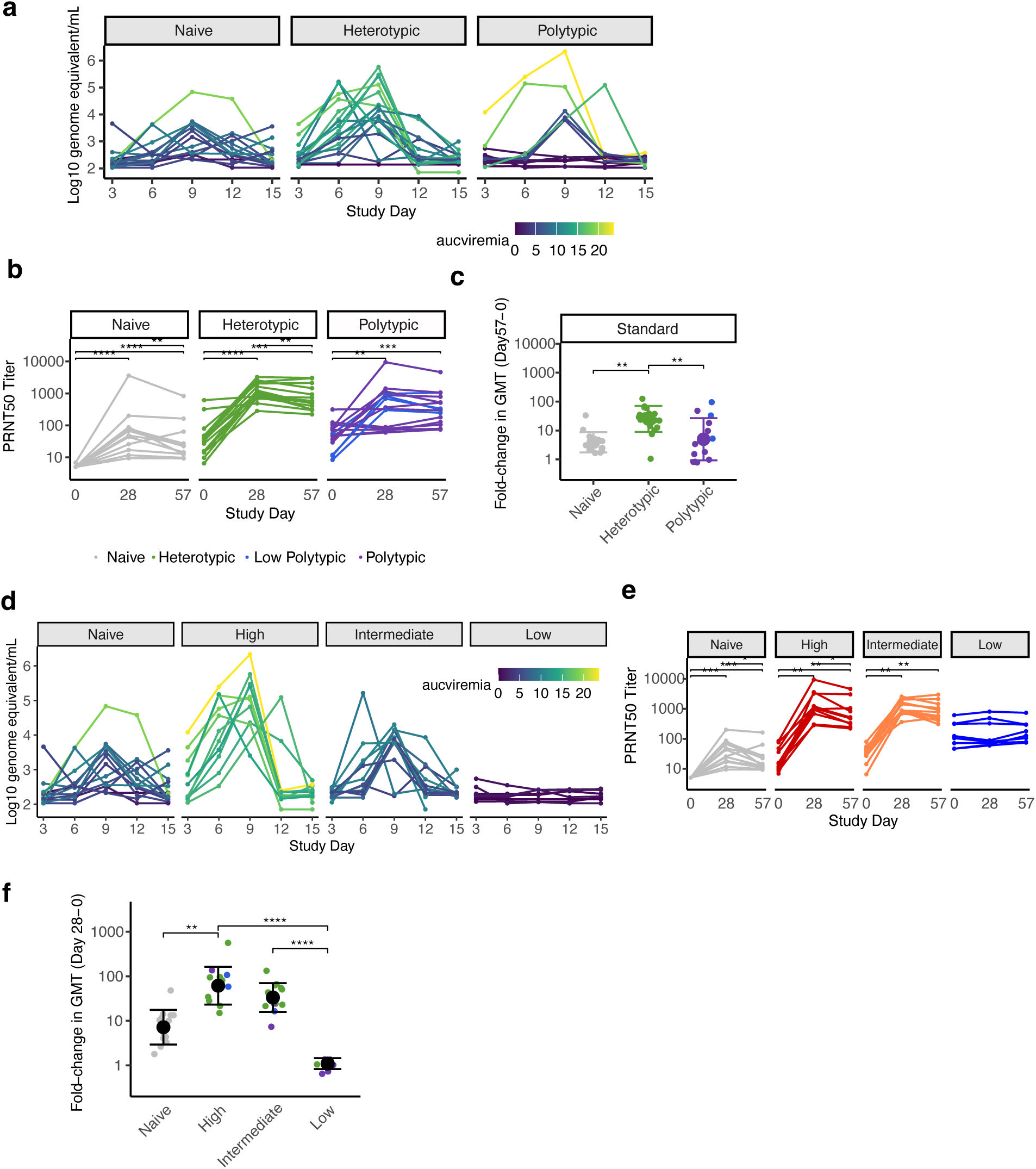
Viremia and DENV1-4 neutralizing antibody geometric mean titers (GMT) by immune group (a-c) and viremia group (d-f). Viremia measured as log_10_ RNA genome equivalent per mL at each timepoint by immune group (a) and viremia group (d). The geometric mean of neutralizing antibodies to standard (partially immature) DENV1-4 by participants and day, stratified by immune group (b) and viremia group (e). Difference in titers at each timepoint within each group was evaluated with unadjusted (as pre-specified for primary endpoints) and adjusted Wilcoxon signed-rank test, respectively. Log_10_ fold-change in geometric mean neutralizing antibodies to standard DENV1-4 by (c) immune group between days 0 and 57 and by (f) viremia group between days 0 and 28. Means and standard deviations are shown. Groups were compared by Kruskal-Wallis test with pairwise comparisons by Dunn’s test with a Bonferroni correction. * p<0.05, ** p<0.01, *** p<0.001, **** p<0.0001.

**Extended Data Fig. 3.**
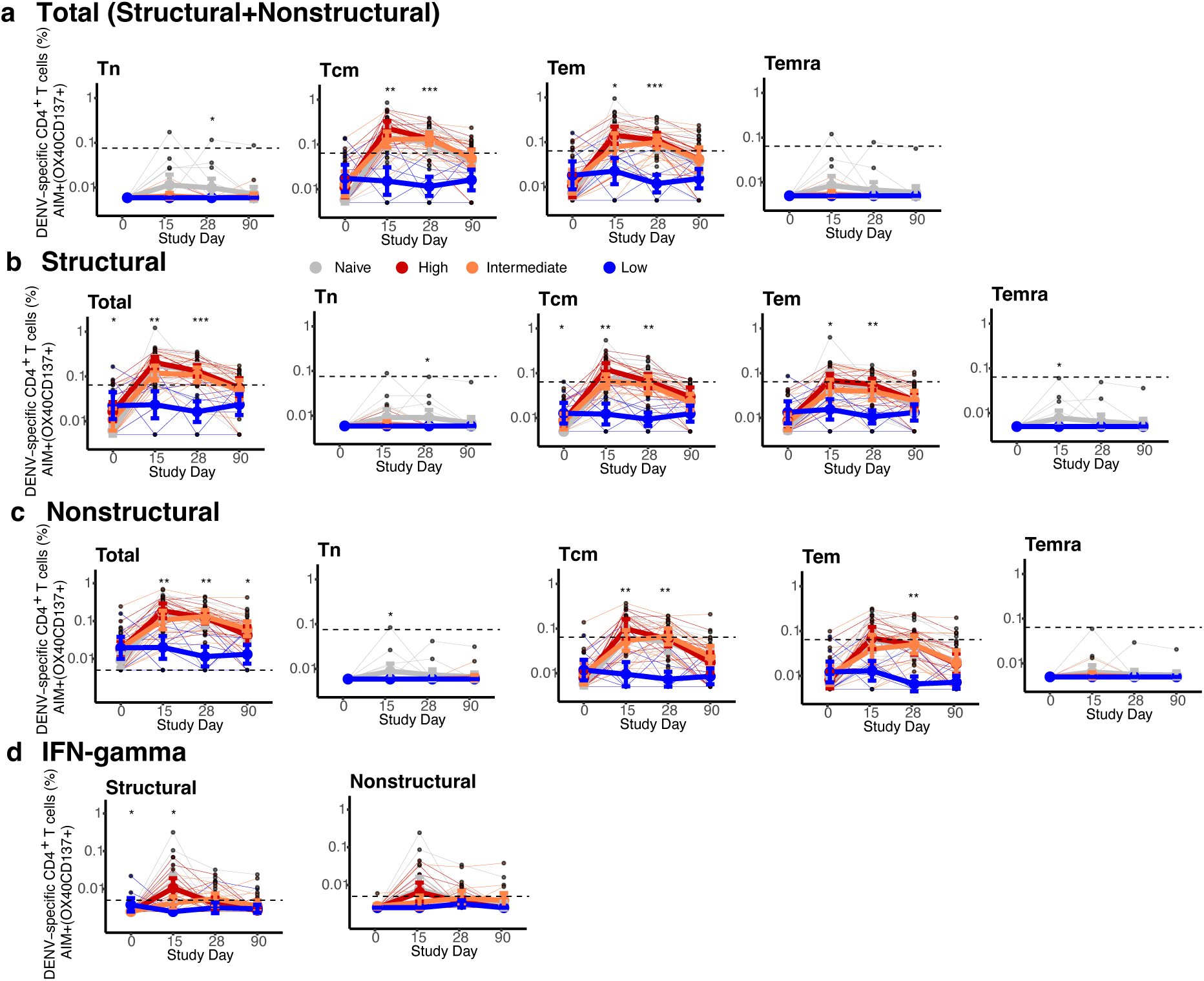
Magnitude of AIM^+^ CD4^+^ T cells across conditions and memory subsets. Figures are stratified by response to (**a**) all DENV proteins, (**b**) only structural proteins (C, prM, E), or (**c**) only non-structural proteins in each viremia group, and (**d**) IFN-γ^+^ CD4^+^ T cells targeting structural or non-structural proteins. Individual data points, viremia group means, and standard deviations shown. Values above the limit of sensitivity (dotted line) are considered responders. Significant differences across groups are indicated with stars and were evaluated by Kruskal-Wallis test. * p<0.05; ** p<0.01; *** p<0.001; **** p<0.0001.

**Extended Data Fig. 4.**
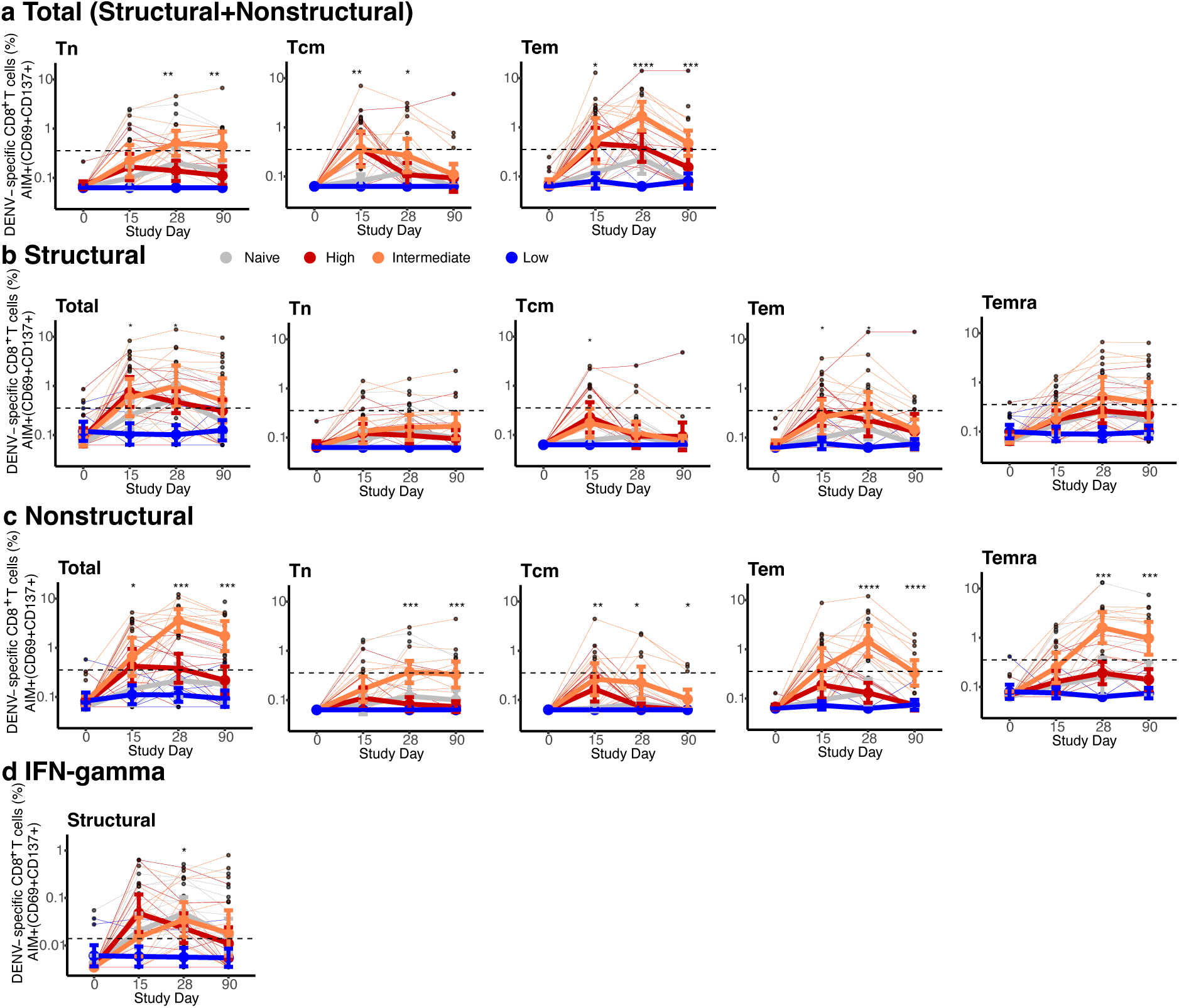
Magnitude of AIM^+^ CD8^+^ T cells across conditions and memory subsets. Figures are stratified by response to (**a**) all DENV proteins, (**b**) only structural proteins (C, prM, E), or (**c**) only non-structural proteins in each viremia group, and (**d**) IFN-γ^+^ CD8^+^ T cells targeting structural proteins. Individual data points, viremia group means, and standard deviations shown. Values above the limit of sensitivity (dotted line) are considered responders. Significant differences across groups are indicated with stars and were evaluated by Kruskal-Wallis test. * p<0.05; ** p<0.01; *** p<0.001; **** p<0.0001.

**Extended Data Fig. 5.**
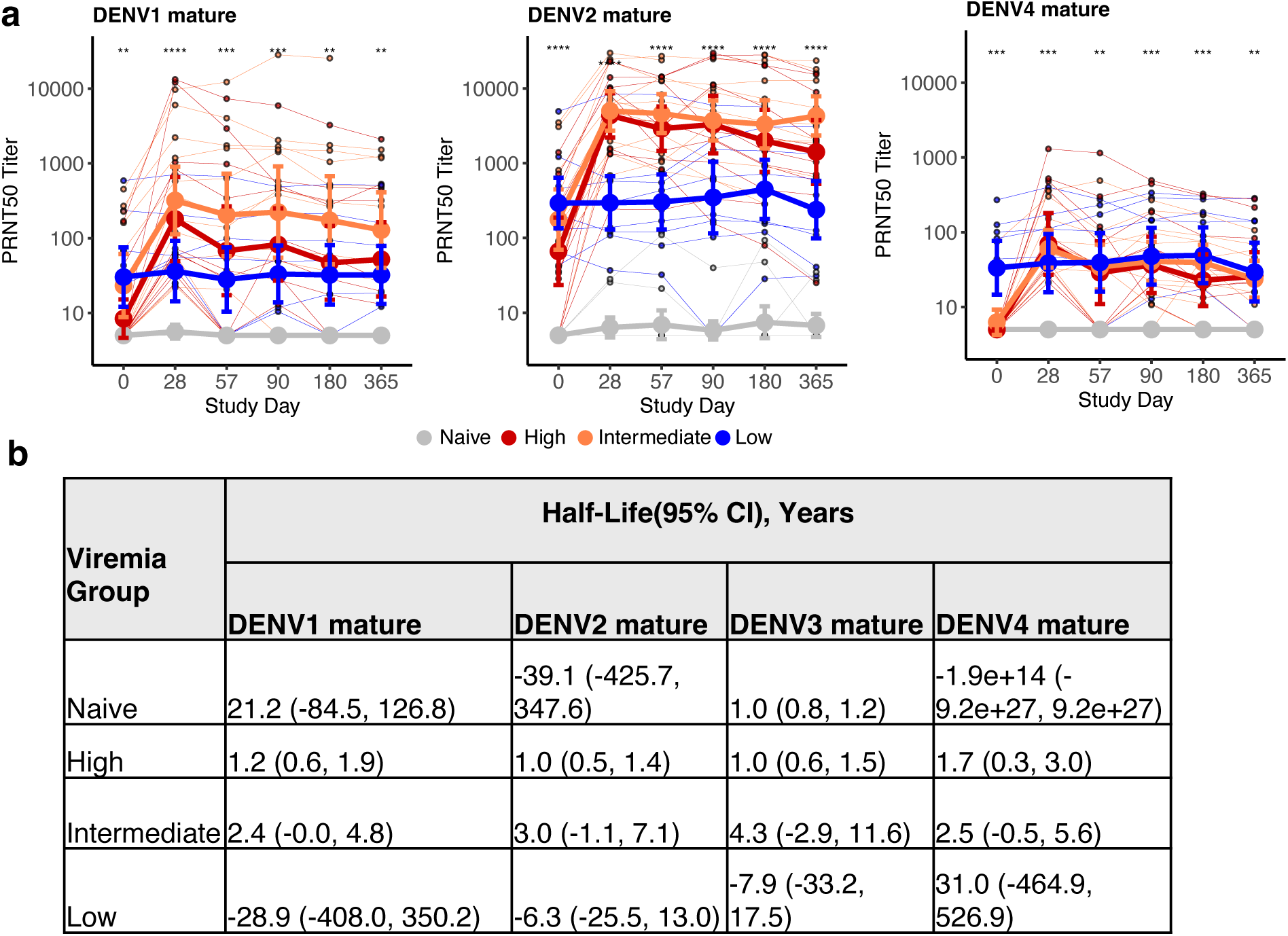
Geometric mean of neutralizing antibodies to (a) mature DENV1, 2, 4 across timepoints, and (b) half-life of mature neutralizing antibody titers to DENV1-4 by viremia group. Across plots, individual datapoints with group means and standard deviations are shown. Titers at each timepoint are compared between viremia groups using Kruskal-Wallis test. * p<0.05; ** p<0.01; *** p<0.001; **** p<0.0001. Half-life and 95% confidence interval (CI) for neutralizing antibodies to DENV1-4 for each viremia group, estimated using a linear mixed-effects model with time-by-group interaction and a subject-specific random intercept to account for repeated measures within individuals.

**Extended Data Fig. 6.**
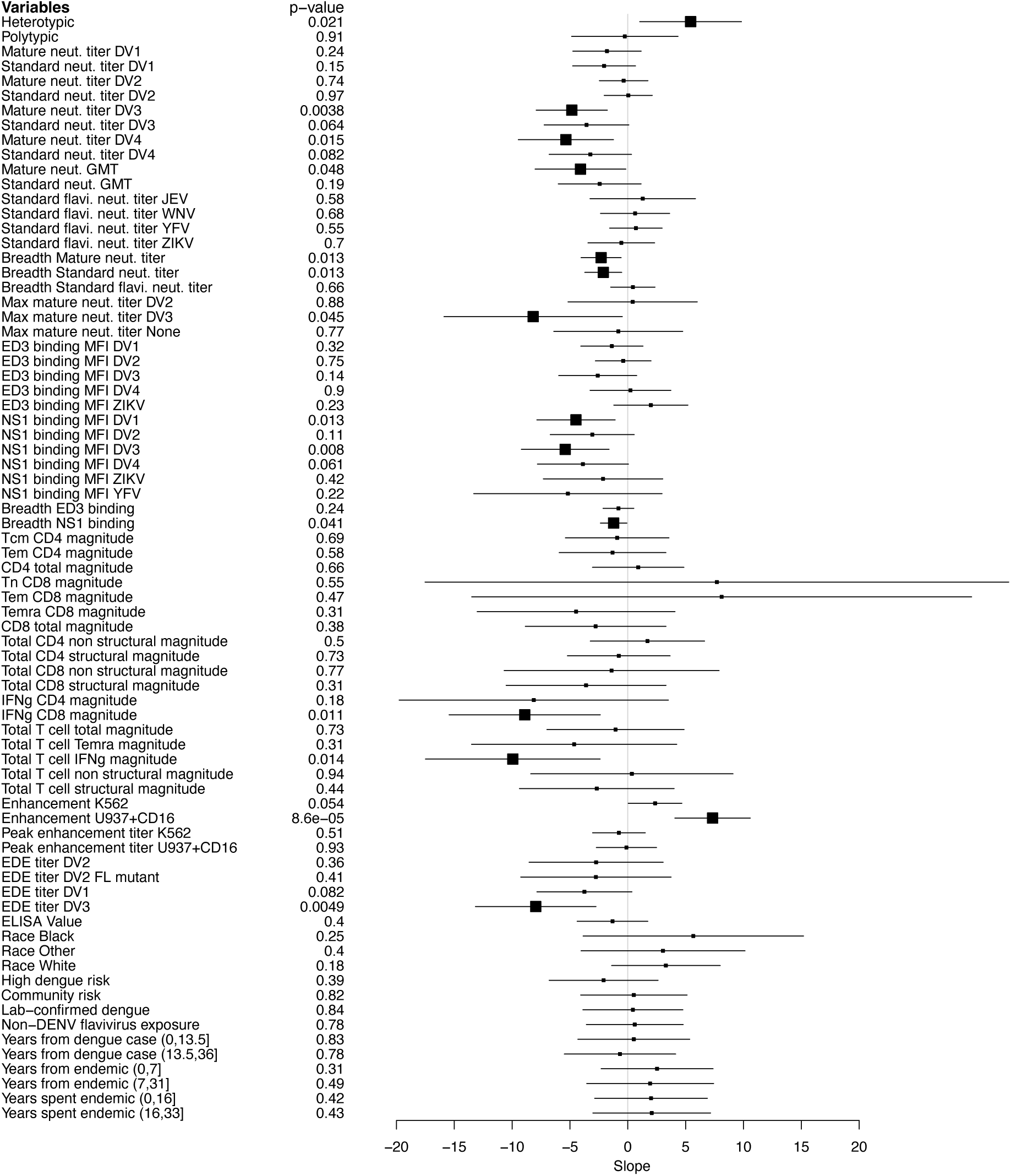
Association between all evaluated baseline immune measures and AUC viremia. Each immune measure was tested individually using a generalized linear model adjusted for age, sex, race, and ethnicity. Slopes with 95% confidence intervals are shown, with effects ordered by related immune and demographic measures.

**Extended Data Fig. 7.**
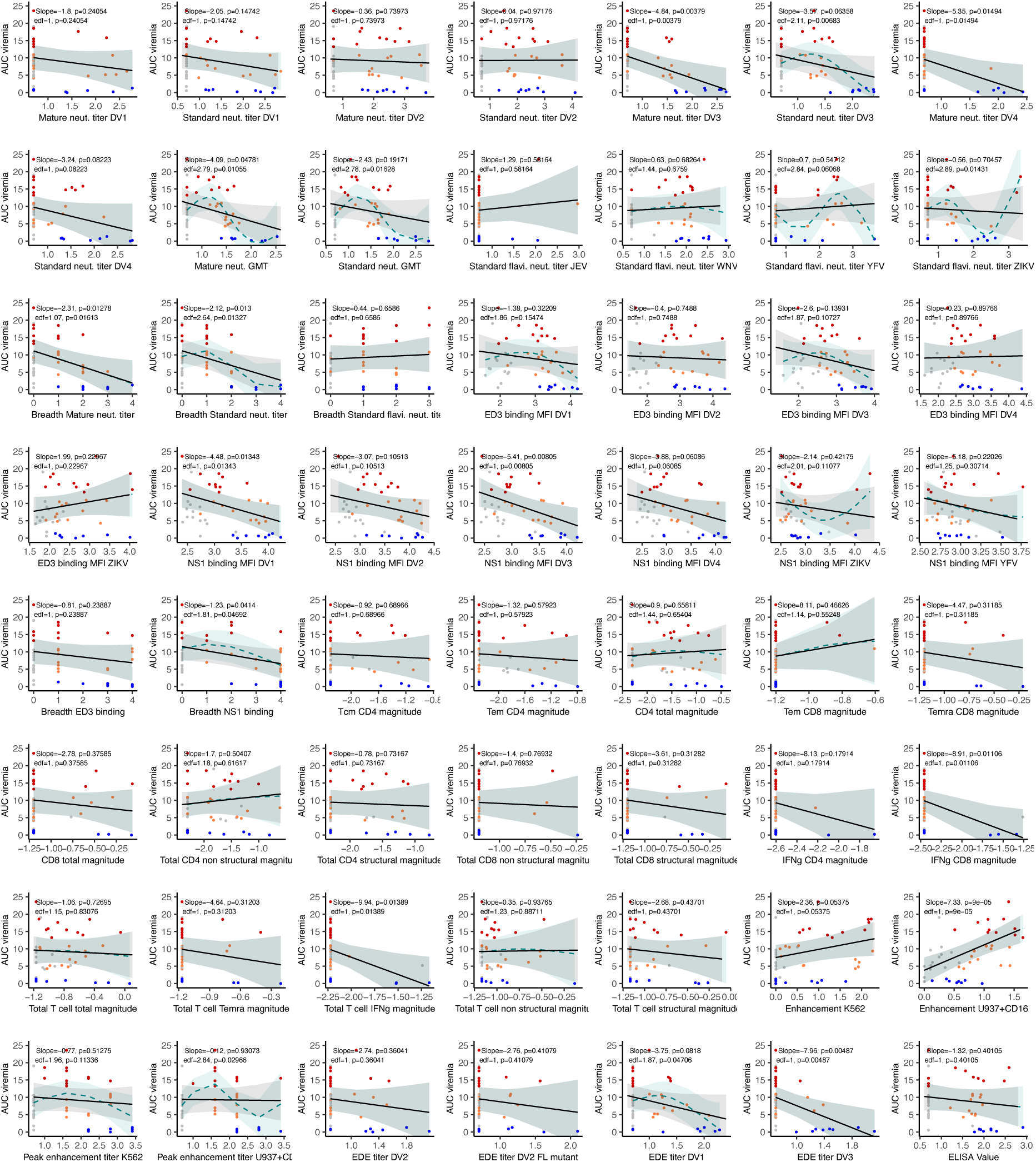
**Linear and non-linear associations between each continuous immune measure and AUC viremia**. Models were adjusted for age, sex, race, and ethnicity. Generalized linear (black) and additive (cyan) model estimates with 95% confidence intervals are shown, along with linear slopes, non-linear estimated degrees of freedom (edf), and associated p-values. Individual datapoints are colored by viremia group.

**Extended Data Fig. 8.**
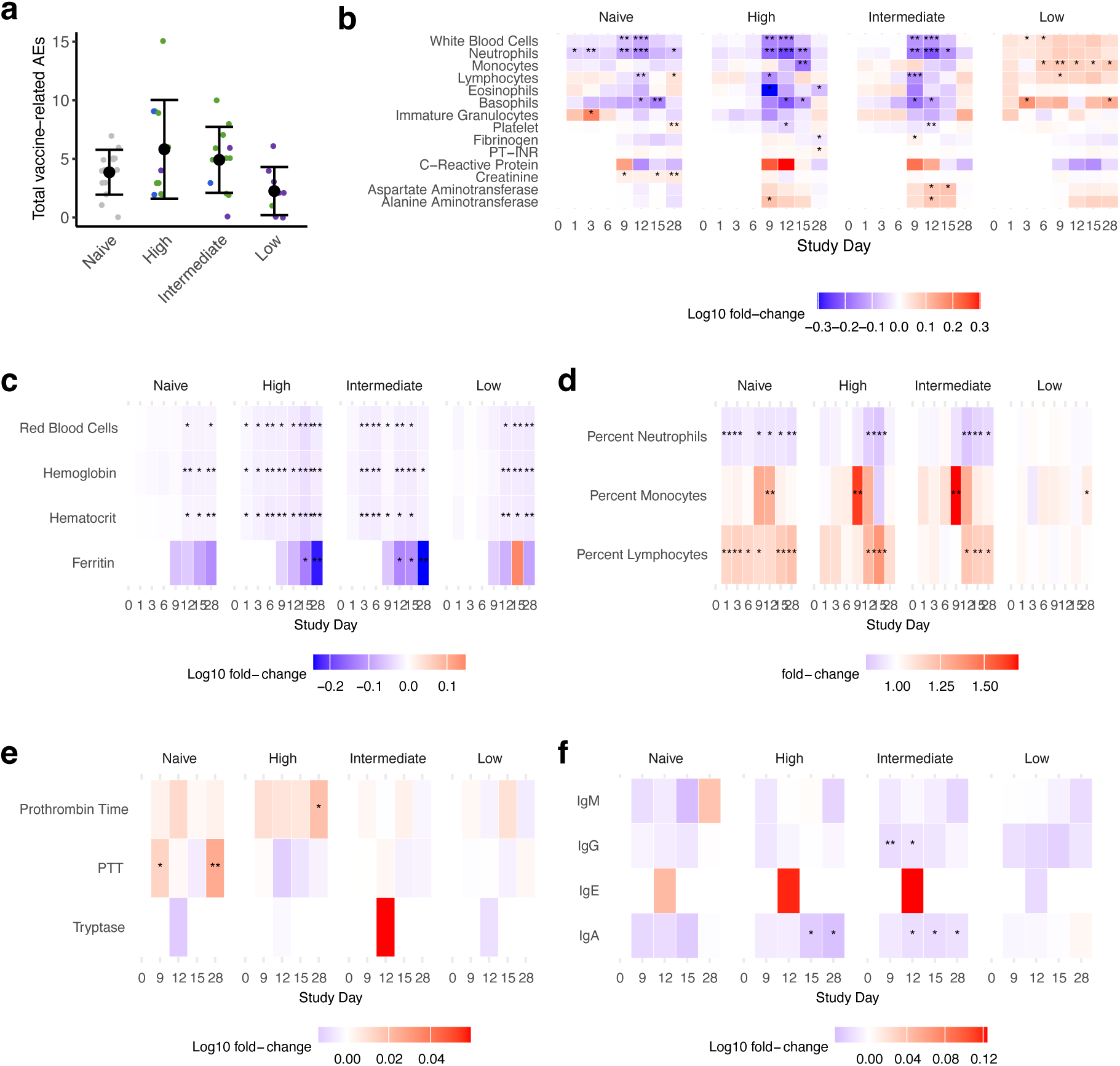
Associations between total vaccine-related adverse events (AEs) and viremia, and heatmaps showing the average log-fold change from day 0 at each timepoint for clinical laboratory values. (a) Total vaccine-related adverse events by viremia group, with means and standard deviations shown. Dots are colored by immune group. Group difference was evaluated by Kruskal-Wallis test but was not significant. The laboratory values assessed included those (b) most commonly affected by DENV infection, (c) measures of red blood cells and iron storage, (d) percentages of notable white blood cell populations, (e) additional coagulation and mast cell markers, and (f) immunoglobulins. Notably, some laboratory values were only collected after a protocol amendment and full data is available for n=28 participants (Fibrinogen, C-Reactive Protein, Tryptase, and all immunoglobulins). Fold-change from day 0 within each viremia group for each laboratory value was evaluated by Wilcoxon signed-rank test. * p<0.05; ** p<0.01; *** p<0.001; **** p<0.0001.

**Extended Data Fig. S9.**
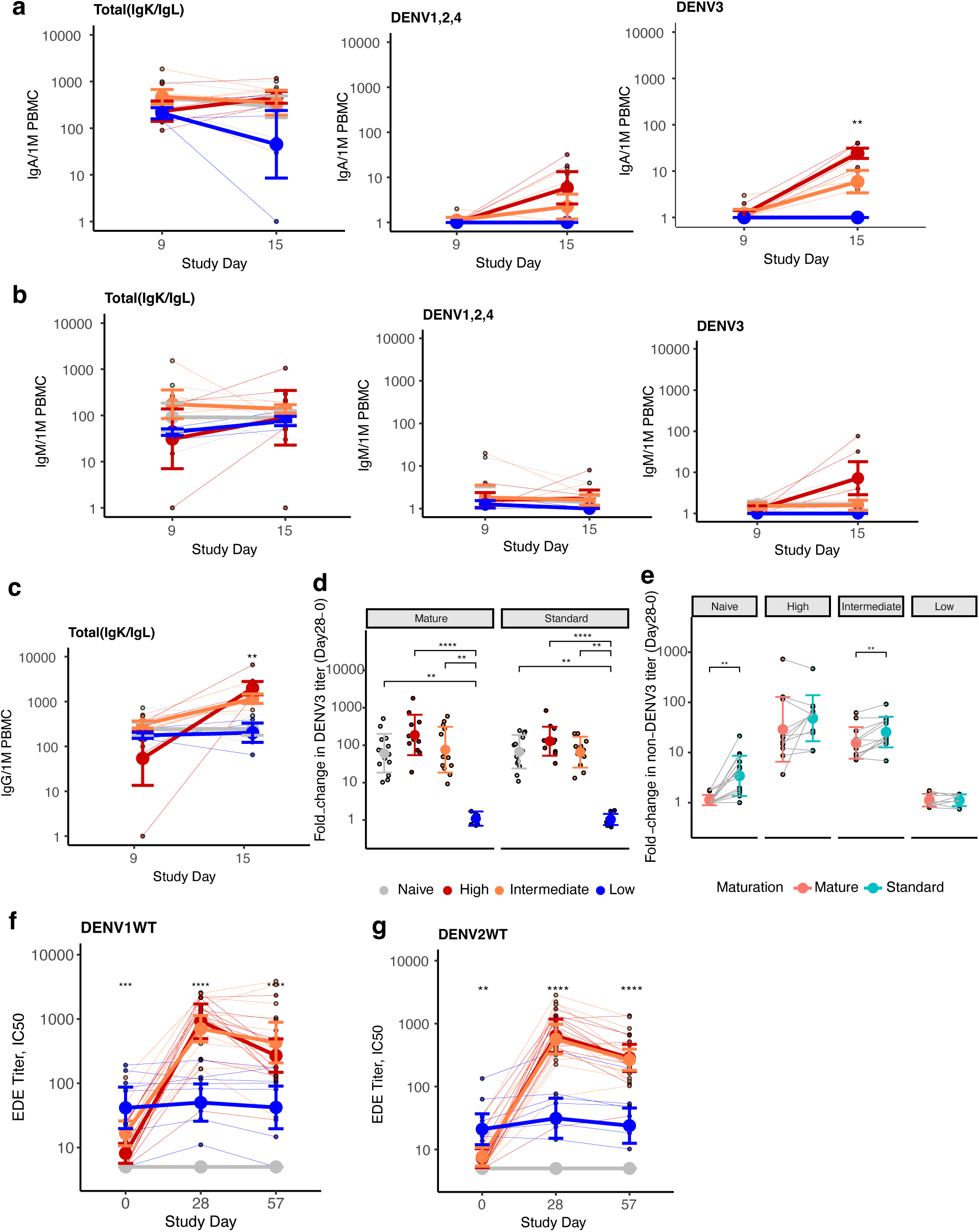
Differences across viremia groups by timepoint in total and DENV-reactive plasmablast populations and changes in neutralizing and functional antibodies over time. Plasmablasts, colored by viremia group, are shown for total plasmablasts per million (1M) PBMCs and those that are reactive to DENV3 or DENV1, 2, and 4 and shown separately for (a) IgA^+^, (b) IgM^+^, and (c) IgG^+^ plasmablasts. A total of 19 participants were evaluated by ELISpot. Comparisons between groups at each timepoint were evaluated by Kruskal-Wallis test. * p<0.05; ** p<0.01; *** p<0.001; *** p<0.0001. **(d)** Fold-change in neutralizing antibodies to mature or standard DENV3 from day 0 to 28 by viremia group. Differences across groups were evaluated by Kruskal-Wallis followed by Dunn’s test with a Bonferroni correction. **(e)** Fold-change from day 0 to 28 in the geometric mean of neutralizing antibodies to standard and mature DENV1, 2, and 4 for each viremia group. Comparisons were evaluated by Wilcoxon signed-rank test. Serum competition with envelope dimer-epitope (EDE) antibody C8 for binding to the DENV1 dimer (**f**) and DENV2 dimer (**g**) at days 0, 28, and 57 for each viremia group. Differences across groups were evaluated by Kruskal-Wallis test. Individual data points are plotted, and group mean and standard deviations shown. * p<0.05; ** p<0.01; *** p<0.001; **** p<0.0001.

**Extended Data Table 1.**
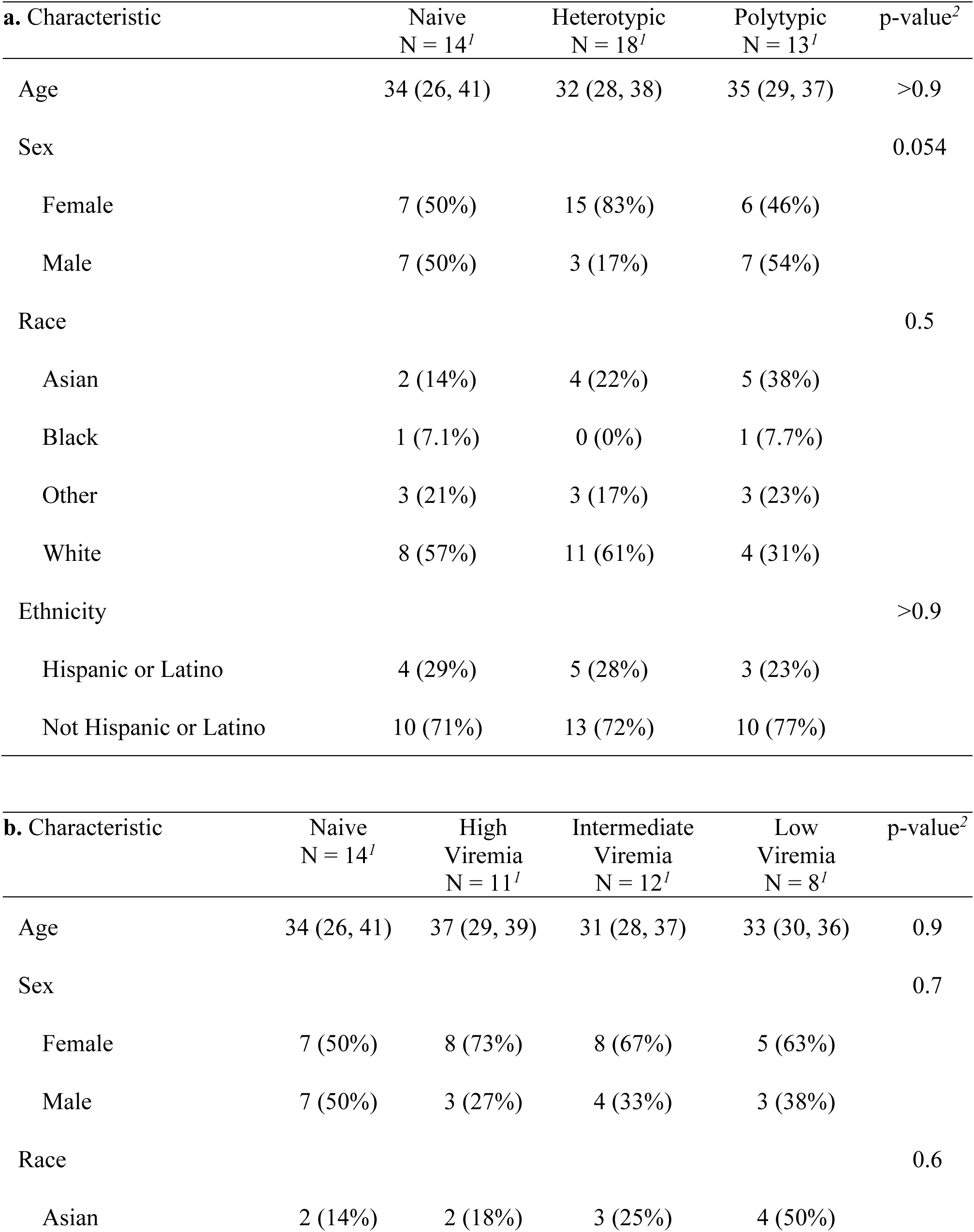
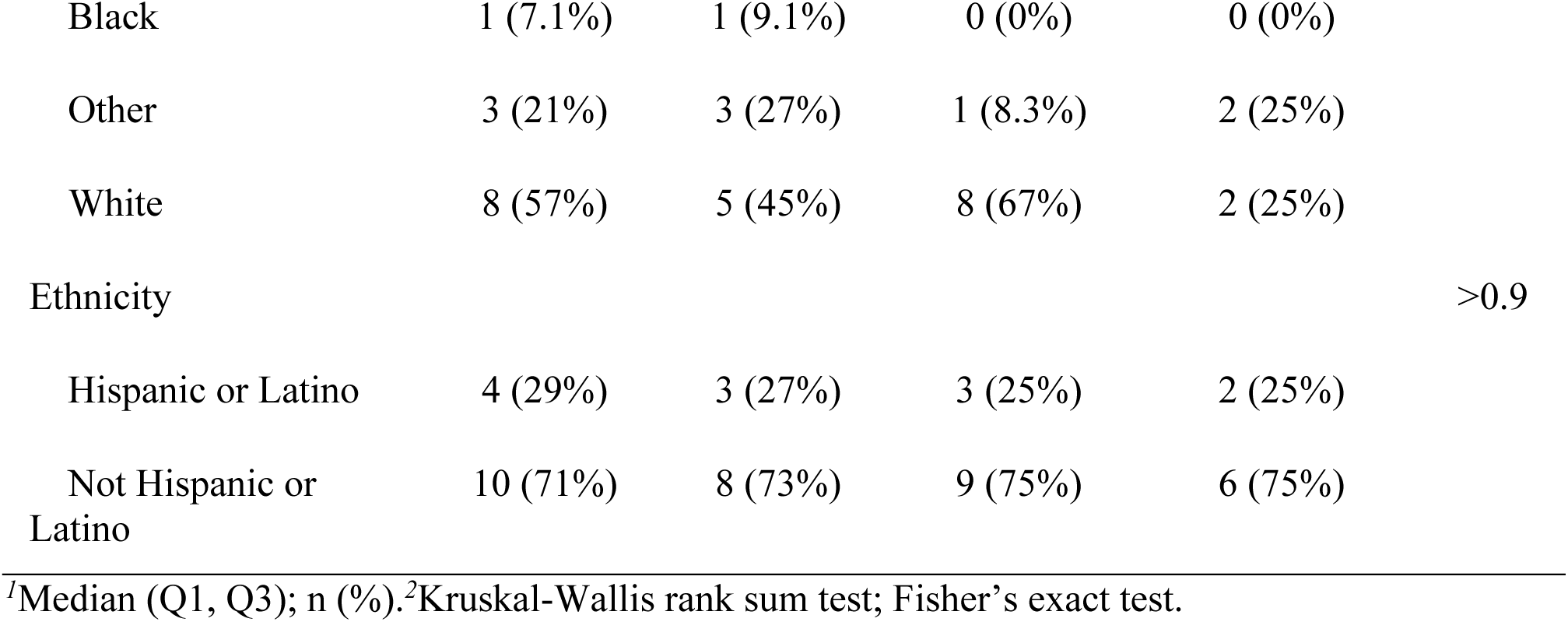
Demographic characteristics for each group. (**a**) By immune group based on day 0 neutralizing antibody titers. (**b**) By viremia group.

## Notes

### Clinical Trial

NCT05691530

### Funding Statement

This study was funded by
Intramural Research Program of the National Institutes of Health
National Institutes of Health Bench to Bedside Program Funds Award 994875
National Institutes of Health Directors Challenge Innovation Award
National Institutes of Health ReVAMPP grant U19AI181960-01
National Institutes of Health P01AI106695
National Cancer Institute NIIH Contract No. 75N91019D00024
The contributions of the NIH authors were made as part of their official duties as NIH federal employees, are in compliance with agency policy requirements, and are considered Works of the United States Government. The findings and conclusions described here are those of the authors and do not necessarily represent the views of the NIH or the U.S. Department of Health and Human Services. In relation to Contract No. 75N91019D00024, the content of this publication does not necessarily reflect the views or policies of the Department of Health and Human Services, nor does mention of trade names, commercial products, or organizations imply endorsement by the U.S. Government.

### Author Declarations

The Institutional Review Board of the National Institutes of Health Office of Human Subjects Research Protection gave ethical approval for this work.

