## Supplementary Information for "Enhancing, controlling, and sterilizing dengue immunity and the development of broadly protective responses"

This PDF file includes: Supplementary Figs. 1-3 and Tables 1-4

Other Supplementary Materials for this manuscript include: Supplementary Data Files 1-4

A

| Reagent | Clone | Source | Catalog # | Dilution / Concentration |
|---|---|---|---|---|
| Golgi-Plug | | BD Biosciences | 555029 | 1:1000 |
| Golgi-Stop | | BD Biosciences | 554724 | 1:1000 |
| LIVE/DEAD Fixable Blue Dead Cell Stain | | Thermo Fisher Scientific | L23105 | 1:50 |
| Human BD Fc Block | | BD Biosciences | 564220 | 1:20 |
| Brilliant Stain Buffer Plus | | BD Biosciences | 566385 | 1:10 |
| CD40 anti-human | HB14 | Miltenyi | 130-094-133 | 0.5 $\mu$g/mL |
| Brilliant Violet 711 anti-human CD197 (CCR7) | G043H7 | BioLegend | 353228 | 1:160 |
| BV605 anti-human CD69 | FN50 | BD Biosciences | 562989 | 1:250 |
| PE-Cy5 anti-human CD137 | 4B4-1 | BioLegend | 309808 | 1:500 |
| BUV805 anti-human CD8 | SK1 | BD Biosciences | 612889 | 1:200 |
| Brilliant Violet 510 anti-human CD16* | 3G8 | BioLegend | 302048 | 1:200 |
| Brilliant Violet 510 anti-human CD14* | 63D3 | BioLegend | 367124 | 1:200 |
| Brilliant Violet 510 anti-human CD20* | 2H7 | BioLegend | 302340 | 1:200 |
| PE-Cy7 anti-human CD134 (OX40) | ACT35 | BD Biosciences | 563663 | 1:160 |
| BUV395 anti-human CD3 | UCHT1 | BD Biosciences | 563546 | 1:100 |
| Brilliant Violet 570 anti-human CD45RA | HI100 | BioLegend | 304132 | 1:100 |
| cFluor BYG750 anti-human CD4 | SK3 | Cytek Biosciences | SKU R7-20159 | 1:100 |
| APC-efluor-780 anti-human CD154 (CD40 Ligand) | 24-31 | Thermo Fisher Scientific | 47-1548-42 | 1:80 |
| BB700 anti-human IFN gamma | B27 | BD Biosciences | 566394 | 1:80 |

*Antibodies make up the dump channel.

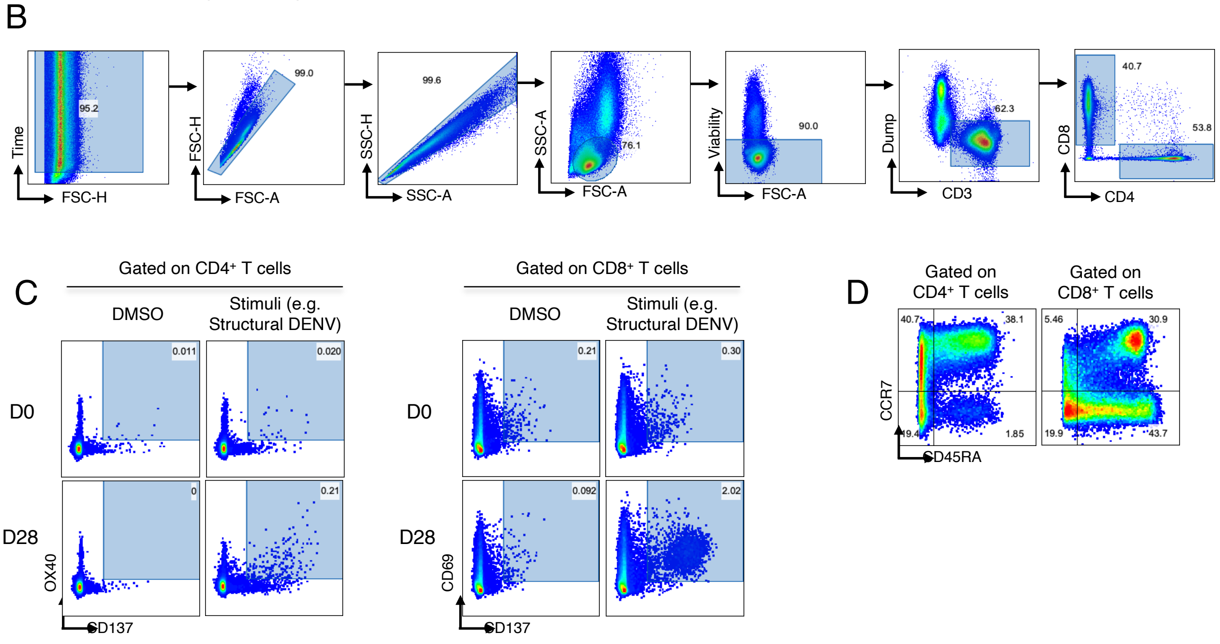


**Supplementary Fig. 1. Representative gating strategy for T cell analysis.** (**A**) Reagents. (**B**) Representative gating strategy to define viable CD4$^+$ and CD8$^+$ lymphocytes. (**C**) Representative gating strategy to define activated induced markers (AIM). (**D**) Representative gating strategy to define memory subsets.

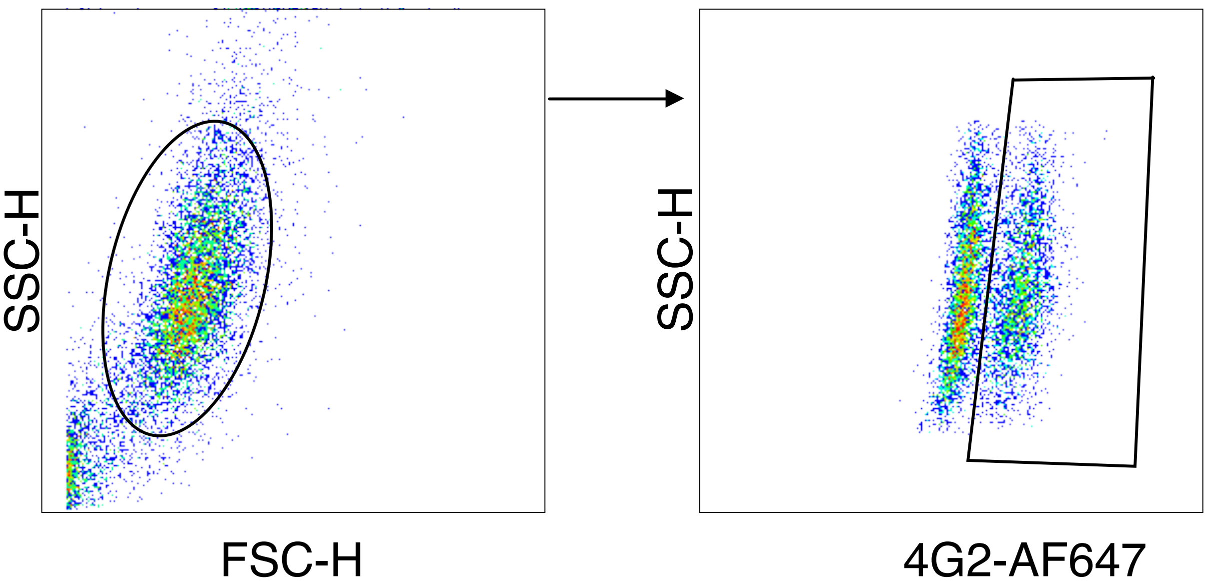


**Supplementary Fig. 2. Representative gating strategy for ADE analysis.** Gating scheme to define DENV-infected U937 cell lines expressing CD16.

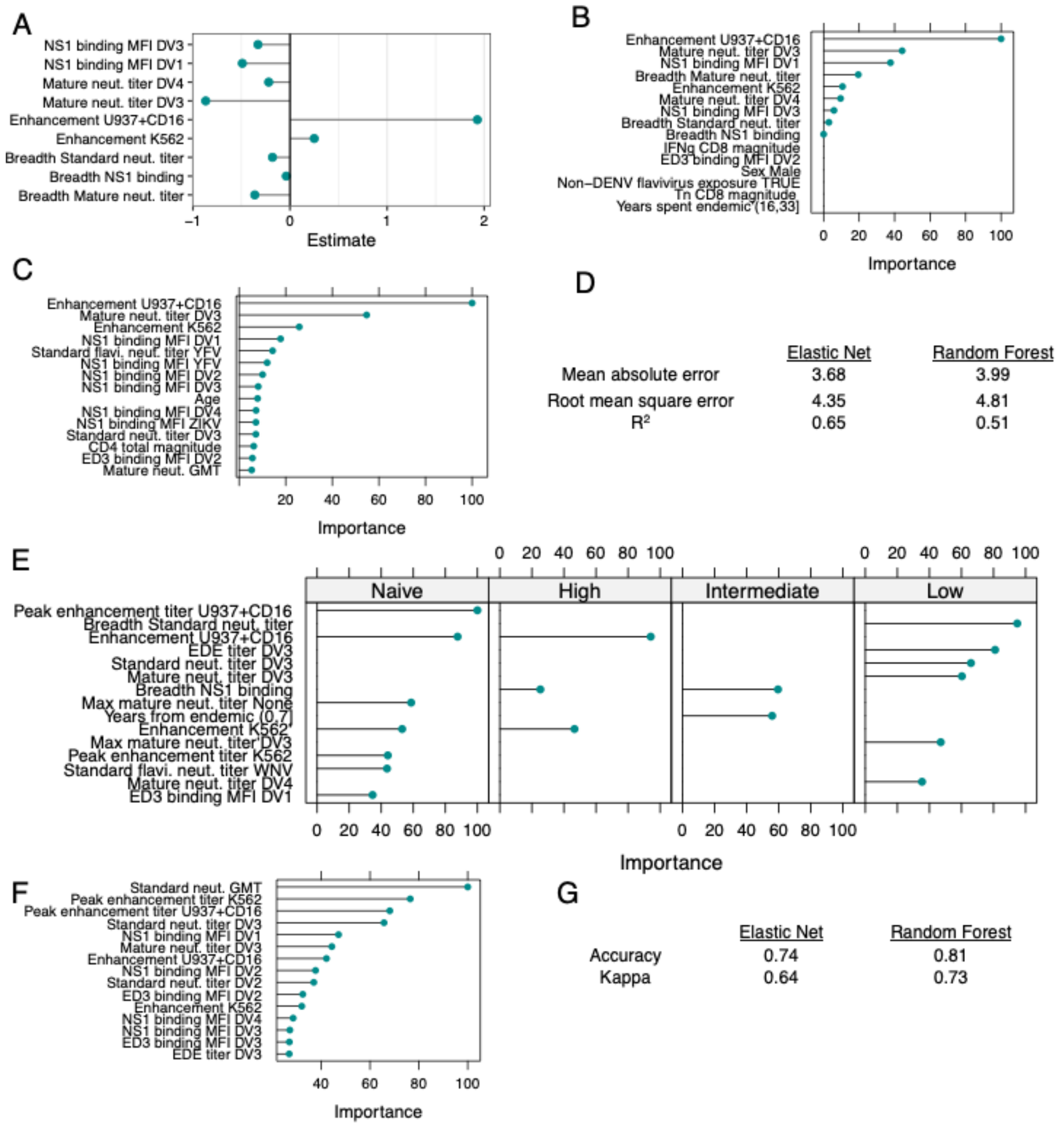


**Supplementary Fig. 3. Immune measures identified as important predictors of viremia using machine learning methods.** Coefficient effect sizes (**A**) and variable importance (**B**) for an elastic net model and variable importance for a random forest (**C**) model evaluating important immune and demographic predictors of AUC viremia. (**D**) Performance of each model was evaluated using cross-validation. Variable importance for elastic net (**E**) and random forest (**F**) multinomial models evaluating important immune and demographic predictors of each viremia group. (**G**) Performance of each model in classifying groups was evaluated using cross-validation.

| Number | Infection history group | DENV1 | DENV2 | DENV3 | DENV4 | GMT |
|---|---|---|---|---|---|---|
| 1 | Naive | 5 | 5 | 5 | 5 | 5 |
| 2 | Naive | 5 | 5 | 5 | 5 | 5 |
| 3 | Naive | 5 | 5 | 5 | 5 | 5 |
| 4 | Naive | 5 | 5 | 5 | 5 | 5 |
| 5 | Naive | 5 | 5 | 5 | 5 | 5 |
| 6 | Naive | 5 | 5 | 5 | 5 | 5 |
| 7 | Naive | 5 | 5 | 5 | 5 | 5 |
| 8 | Naive | 5 | 5 | 5 | 5 | 5 |
| 9 | Naive | 5 | 5 | 5 | 5 | 5 |
| 10 | Naive | 5 | 5 | 5 | 5 | 5 |
| 11 | Naive | 5 | 5 | 5 | 5 | 5 |
| 12 | Naive | 5 | 5 | 5 | 5 | 5 |
| 13 | Naive | 5 | 5 | 5 | 5 | 5 |
| 14 | Naive | 5 | 5 | 5 | 5 | 5 |
| 15 | Heterotypic | 14 | 5 | 5 | 5 | 6 |
| 16 | Heterotypic | 18 | 5 | 5 | 5 | 7 |
| 17 | Heterotypic | 5 | 39 | 5 | 5 | 8 |
| 18 | Heterotypic | 5 | 71 | 5 | 5 | 10 |
| 19 | Heterotypic | 5 | 332 | 5 | 5 | 14 |

| | | | | | | |
|---|---|---|---|---|---|---|
| 20 | Heterotypic | 5 | 375 | 5 | 5 | 15 |
| 21 | Heterotypic | 91 | 11 | 12 | 5 | 16 |
| 22 | Heterotypic | 10 | 642 | 17 | 5 | 27 |
| 23 | Heterotypic | 546 | 78 | 17 | 5 | 29 |
| 24 | Heterotypic | 5 | 4302 | 11 | 5 | 33 |
| 25 | Heterotypic | 5 | 371 | 28 | 33 | 36 |
| 26 | Heterotypic | 415 | 85 | 14 | 5 | 39 |
| 27 | Heterotypic | 11 | 507 | 27 | 24 | 43 |
| 28 | Heterotypic | 727 | 44 | 27 | 5 | 46 |
| 29 | Heterotypic | 24 | 429 | 21 | 24 | 48 |
| 30 | Heterotypic | 15 | 1302 | 43 | 39 | 76 |
| 31 | Heterotypic | 15 | 4287 | 29 | 21 | 79 |
| 32 | Heterotypic | 475 | 12812 | 39 | 592 | 614 |
| 33 | Polytypic low | 5 | 32 | 20 | 5 | 11 |
| 34 | Polytypic low | 5 | 42 | 20 | 5 | 12 |
| 35 | Polytypic low | 23 | 51 | 50 | 161 | 55 |
| 36 | Polytypic | 101 | 76 | 22 | 10 | 36 |
| 37 | Polytypic | 16 | 21 | 102 | 133 | 47 |
| 38 | Polytypic | 14 | 136 | 129 | 19 | 47 |
| 39 | Polytypic | 145 | 82 | 32 | 12 | 47 |
| 40 | Polytypic | 20 | 265 | 241 | 20 | 71 |
| 41 | Polytypic | 208 | 115 | 37 | 57 | 85 |

| | | | | | | |
|---|---|---|---|---|---|---|
| 42 | Polytypic | 194 | 207 | 135 | 22 | 105 |
| 43 | Polytypic | 251 | 112 | 100 | 83 | 123 |
| 44 | Polytypic | 529 | 558 | 177 | 187 | 314 |
| 45 | Polytypic | 73 | 896 | 245 | 658 | 320 |

**Supplementary Table 1. Day 0 neutralizing antibody titers for all participants.**

| Characteristic | Naive<br>N = 14[1] | Heterotypic<br>N = 18[1] | Polytypic<br>N = 13[1] | p-value[2] |
|---|---|---|---|---|
| Yellow fever vaccination | | | | 0.001 |
| No | 14 (100%) | 7 (39%) | 6 (46%) | |
| Not sure | 0 (0%) | 4 (22%) | 1 (7.7%) | |
| Yes | 0 (0%) | 7 (39%) | 6 (46%) | |
| Japanese encephalitis vaccination | | | | 0.2 |
| No | 14 (100%) | 13 (72%) | 11 (85%) | |
| Not sure | 0 (0%) | 4 (22%) | 1 (7.7%) | |
| Yes | 0 (0%) | 1 (5.6%) | 1 (7.7%) | |
| Tick-borne encephalitis vaccination | | | | 0.14 |
| No | 14 (100%) | 14 (78%) | 10 (77%) | |
| Not sure | 0 (0%) | 4 (22%) | 2 (15%) | |
| Yes | 0 (0%) | 0 (0%) | 1 (7.7%) | |
| Zika case | 0 (0%) | 1 (5.6%) | 0 (0%) | >0.9 |
| Dengue case | 0 (0%) | 12 (67%) | 10 (77%) | <0.001 |
| Lab-confirmed dengue case | 0 (0%) | 7 (39%) | 6 (46%) | 0.007 |
| Years since dengue case | NA | 13 (6, 17) | 15 (9, 18) | 0.6 |

| | | | | |
|---|---|---|---|---|
| Not applicable (n) | 14 | 6 | 3 | |
| Years since living in endemic area | 5 (4, 12) | 7 (4, 10) | 7 (3, 15) | 0.9 |
| Not applicable (n) | 9 | 1 | 2 | |
| Total time living in endemic area | 21 (8, 28) | 11 (4, 26) | 19 (5, 23) | >0.9 |
| Not applicable (n) | 10 | 2 | 4 | |
| Contacts with dengue exposure | 2 (14%) | 4 (22%) | 8 (62%) | 0.022 |
| Community took preventative measures against dengue | 4 (29%) | 12 (67%) | 9 (69%) | 0.049 |

[1]Median (Q1, Q3); n (%).

[2]Kruskal-Wallis rank sum test; Fisher's exact test.

**Supplementary Table 2. History of flavivirus vaccination, infection, and exposure risk.**

| Adverse Event | Naïve (N=14) | Heterotypic (N=18) | Polytypic (N=13) | p-value[1] |
|---|---|---|---|---|
| Dengue-Like Rash | 78.6 | 44.4 | 23.1 | 0.015 |
| Feverish (subjective) | 0 | 0 | 23.1 | 0.078 |
| Diarrhea (>1 loose stool/day) | 0 | 22.2 | 0 | 0.097 |
| Myalgia | 7.1 | 38.9 | 15.4 | 0.107 |
| Respiratory rate >16 breaths per minute | 21.4 | 27.8 | 0 | 0.117 |
| Diastolic blood pressure >90 mmHg | 14.3 | 0 | 23.1 | 0.124 |
| Aspiration site bruising | 21.4 | 22.2 | 0 | 0.182 |
| Arthralgia | 0 | 22.2 | 7.7 | 0.187 |
| Headache | 57.1 | 27.8 | 30.8 | 0.22 |
| Glucose >99 (fasting) or >109 (random) mg/dL | 50 | 22.2 | 38.5 | 0.27 |
| Venipuncture site bruise | 14.3 | 0 | 0 | 0.271 |
| Hemoglobin <12.6 (male) or 11.1 (female) g/dL | 21.4 | 44.4 | 23.1 | 0.33 |
| Neutrophils <1,500 K/uL | 21.4 | 44.4 | 23.1 | 0.33 |
| Bruising | 0 | 16.7 | 7.7 | 0.349 |
| Fibrinogen >399 mg/dL | 0 | 5.6 | 15.4 | 0.396 |
| Acid reflux (esophageal) | 0 | 11.1 | 0 | 0.456 |
| Increased thirst | 0 | 11.1 | 0 | 0.456 |
| Injection site pain | 0 | 11.1 | 0 | 0.456 |
| Photophobia | 0 | 11.1 | 0 | 0.456 |

| | | | | |
|---|---|---|---|---|
| Injection site tenderness | 0 | 11.1 | 0 | 0.456 |
| Potassium <3.7 mmol/L | 28.6 | 11.1 | 23.1 | 0.458 |
| Heart rate >100 beats per minute | 14.3 | 11.1 | 0 | 0.461 |
| Lymphocyte count <1,001K/uL | 7.1 | 22.2 | 7.7 | 0.468 |
| Malaise | 7.1 | 22.2 | 7.7 | 0.468 |
| Rash | 7.1 | 22.2 | 7.7 | 0.468 |
| Leukocyte count <3,501 K/mcL | 21.4 | 38.9 | 23.1 | 0.528 |
| Systolic blood pressure >140 mmHg | 7.1 | 0 | 7.7 | 0.561 |
| Hemoglobin drop >1.5 gm/dL from baseline | 0 | 11.1 | 7.7 | 0.562 |
| Retro-orbital pain | 0 | 11.1 | 7.7 | 0.562 |
| Liver enzymes >1.0x upper limit normal (ULN) | 0 | 0 | 7.7 | 0.577 |
| Hypersensitivity | 0 | 0 | 7.7 | 0.577 |
| Paresthesia | 0 | 0 | 7.7 | 0.577 |
| Polydipsia | 0 | 0 | 7.7 | 0.577 |
| BUN >22 mg/dL | 0 | 0 | 7.7 | 0.577 |
| C-reactive protein >5 mg/L | 0 | 0 | 7.7 | 0.577 |
| Insect bite | 0 | 0 | 7.7 | 0.577 |
| Shortness of breath | 0 | 0 | 7.7 | 0.577 |
| Sunburn-like rash | 0 | 0 | 7.7 | 0.577 |
| Tenderness | 0 | 0 | 7.7 | 0.577 |
| Vasovagal reaction | 7.1 | 11.1 | 0 | 0.605 |

| | | | | |
|---|---|---|---|---|
| Creatinine kinase >1.24x ULN | 7.1 | 0 | 0 | 0.622 |
| Dry skin | 7.1 | 0 | 0 | 0.622 |
| Dysuria | 7.1 | 0 | 0 | 0.622 |
| Hoarseness | 7.1 | 0 | 0 | 0.622 |
| Cold sores | 7.1 | 0 | 0 | 0.622 |
| Cough | 7.1 | 0 | 0 | 0.622 |
| Joint pain | 7.1 | 0 | 0 | 0.622 |
| Nasal congestion | 7.1 | 0 | 0 | 0.622 |
| Presyncope | 7.1 | 0 | 0 | 0.622 |
| Venipuncture site pain | 7.1 | 0 | 0 | 0.622 |
| Fatigue | 57.1 | 50 | 38.5 | 0.639 |
| Chills | 7.1 | 16.7 | 7.7 | 0.73 |
| AST >1x ULN | 14.3 | 5.6 | 7.7 | 0.73 |
| Sore throat | 14.3 | 5.6 | 7.7 | 0.73 |
| Aspiration site pain | 21.4 | 33.3 | 23.1 | 0.748 |
| Injection site redness | 7.1 | 5.6 | 0 | 0.777 |
| ALT >1x ULN | 7.1 | 5.6 | 0 | 0.777 |
| Viral infection NOS | 7.1 | 5.6 | 0 | 0.777 |
| Abdominal cramps | 0 | 5.6 | 0 | 0.799 |
| Aspiration site swelling | 0 | 5.6 | 0 | 0.799 |
| Constipation | 0 | 5.6 | 0 | 0.799 |
| Decreased appetite | 0 | 5.6 | 0 | 0.799 |
| Difficulty sleeping | 0 | 5.6 | 0 | 0.799 |

| | | | | |
|---|---|---|---|---|
| Dizziness | 0 | 5.6 | 0 | 0.799 |
| Drenching sweats | 0 | 5.6 | 0 | 0.799 |
| Eye pain | 0 | 5.6 | 0 | 0.799 |
| Fall | 0 | 5.6 | 0 | 0.799 |
| Herpes simplex type I | 0 | 5.6 | 0 | 0.799 |
| Potassium >5 mEq/L | 0 | 5.6 | 0 | 0.799 |
| Systolic blood pressure <90 mmHg | 0 | 5.6 | 0 | 0.799 |
| Injection site erythema | 0 | 5.6 | 0 | 0.799 |
| Low back pain | 0 | 5.6 | 0 | 0.799 |
| Night sweats | 0 | 5.6 | 0 | 0.799 |
| PTT >1x ULN | 0 | 5.6 | 0 | 0.799 |
| Periorbital edema | 0 | 5.6 | 0 | 0.799 |
| Pneumonia | 0 | 5.6 | 0 | 0.799 |
| Sinus congestion | 0 | 5.6 | 0 | 0.799 |
| Urinary frequency | 0 | 5.6 | 0 | 0.799 |
| Urinary tract infection | 0 | 5.6 | 0 | 0.799 |
| Glucose <70 mg/dL | 14.3 | 16.7 | 7.7 | 0.818 |
| Lymphadenopathy | 14.3 | 11.1 | 7.7 | 0.895 |
| Heart rate <55 beats per minute | 7.1 | 11.1 | 7.7 | 0.955 |
| Sodium <135 mEq/L | 7.1 | 5.6 | 7.7 | 0.958 |
| Nausea | 14.3 | 16.7 | 15.4 | 0.991 |

[1]p-value for likelihood ratio test, overall differences among groups.

**Supplementary Table 3. Frequency (%) of all adverse events through day 28 across immune groups based on day 0 neutralizing antibody titers.**

| Adverse Event | Naïve (N=14) | Heterotypic (N=18) | Polytypic (N=13) | p-value[1] |
|---|---|---|---|---|
| Dengue-Like Rash | 78.6 | 44.4 | 23.1 | 0.015 |
| Feverish (subjective) | 0 | 0 | 23.1 | 0.078 |
| Myalgia | 7.1 | 33.3 | 15.4 | 0.209 |
| Diarrhea (>1 loose stool/day) | 0 | 16.7 | 0 | 0.221 |
| Diastolic blood pressure >90 mmHg | 14.3 | 0 | 0 | 0.271 |
| Headache | 50 | 27.8 | 23.1 | 0.306 |
| Neutrophils <1,500 K/uL | 21.4 | 44.4 | 23.1 | 0.33 |
| Leukocyte count <3,501 K/mcL | 21.4 | 38.9 | 15.4 | 0.348 |
| Arthralgia | 0 | 16.7 | 7.7 | 0.349 |
| Increased thirst | 0 | 11.1 | 0 | 0.456 |
| Injection site pain | 0 | 11.1 | 0 | 0.456 |
| Photophobia | 0 | 11.1 | 0 | 0.456 |
| Injection site tenderness | 0 | 11.1 | 0 | 0.456 |
| Lymphocyte count <1,001K/uL | 7.1 | 22.2 | 7.7 | 0.468 |
| Malaise | 7.1 | 22.2 | 7.7 | 0.468 |
| Retro-orbital pain | 0 | 11.1 | 7.7 | 0.562 |

| | | | | |
|---|---|---|---|---|
| Liver enzymes >1.0x upper limit normal (ULN) | 0 | 0 | 7.7 | 0.577 |
| Hypersensitivity | 0 | 0 | 7.7 | 0.577 |
| Paresthesia | 0 | 0 | 7.7 | 0.577 |
| Polydipsia | 0 | 0 | 7.7 | 0.577 |
| C-reactive protein >5 mg/L | 0 | 0 | 7.7 | 0.577 |
| Shortness of breath | 0 | 0 | 7.7 | 0.577 |
| Tenderness | 0 | 0 | 7.7 | 0.577 |
| Rash | 7.1 | 11.1 | 0 | 0.605 |
| Joint pain | 7.1 | 0 | 0 | 0.622 |
| Systolic blood pressure >140 mmHg | 7.1 | 0 | 0 | 0.622 |
| Vasovagal reaction | 7.1 | 0 | 0 | 0.622 |
| Fatigue | 57.1 | 50 | 38.5 | 0.639 |
| Bruising | 0 | 5.6 | 7.7 | 0.721 |
| Chills | 7.1 | 16.7 | 7.7 | 0.73 |
| Lymphadenopathy | 14.3 | 5.6 | 7.7 | 0.73 |
| Injection site redness | 7.1 | 5.6 | 0 | 0.777 |
| ALT >1x ULN | 0 | 5.6 | 0 | 0.799 |
| Aspiration site swelling | 0 | 5.6 | 0 | 0.799 |
| Decreased appetite | 0 | 5.6 | 0 | 0.799 |
| Difficulty sleeping | 0 | 5.6 | 0 | 0.799 |
| Dizziness | 0 | 5.6 | 0 | 0.799 |

| | | | | |
|---|---|---|---|---|
| Drenching sweats | 0 | 5.6 | 0 | 0.799 |
| Injection site erythema | 0 | 5.6 | 0 | 0.799 |
| Low back pain | 0 | 5.6 | 0 | 0.799 |
| Night sweats | 0 | 5.6 | 0 | 0.799 |
| PTT >1x ULN | 0 | 5.6 | 0 | 0.799 |
| Periorbital edema | 0 | 5.6 | 0 | 0.799 |
| Pruritus | 0 | 5.6 | 0 | 0.799 |
| Heart rate >100 beats per minute | 0 | 5.6 | 0 | 0.799 |
| Nausea | 14.3 | 11.1 | 15.4 | 0.925 |
| AST >1x ULN | 7.1 | 5.6 | 7.7 | 0.958 |

[1]p-value for likelihood ratio test, overall differences among groups

**Supplementary Table 4. Frequency (%) of vaccine-associated adverse events across immune groups based on day 0 neutralizing antibody titers.**
